# The military as a neglected pathogen transmitter, from the 19th century to COVID-19: A systematic review

**DOI:** 10.1101/2021.10.09.21264758

**Authors:** Claudia Chaufan, Ilinca A. Dutescu, Hanah Fekre, Saba Marzabadi, K.J. Noh

## Abstract

**Background:** The risk of outbreaks escalating into pandemics has soared with globalization. Therefore, understanding transmission mechanisms of infectious diseases has become critical to formulating global public health policy. This systematic review assessed evidence in the medical and public health literature for the military as a disease vector.

**Methods:** We searched 3 electronic databases without temporal restrictions. Two researchers independently extracted study data using a standardized form. Through team discussions, studies were grouped according to their type of transmission mechanism and direct quotes were extracted to generate themes and sub-themes. A content analysis was later performed and frequency distributions for each theme were generated.

**Results:** Of 6477 studies, 210 met our inclusion criteria and provided evidence, spanning over two centuries (1810 – 2020), for the military as a pathogen transmitter, within itself or between it and civilians. Biological mechanisms driving transmission included person-to-person transmission, contaminated food and water, vector-borne, and airborne routes. Contaminated food and/or water were the most common biological transmission route. Social mechanisms facilitating transmission included crowded living spaces, unhygienic conditions, strenuous working, training conditions, absent or inadequate vaccination programs, pressure from military leadership, poor compliance with public health advice, contractor mismanagement, high-risk behaviours, and occupation-specific freedom of movement. Living conditions were the most common social transmission mechanism, with young, low ranking military personnel repeatedly reported as the most affected group. Selected social mechanisms, such as employment-related freedom of movement, were unique to the military as a social institution. While few studies explicitly studied civilian populations, considerably more contained information that implied that civilians were likely impacted by outbreaks described in the military.

**Conclusions:** This study identified features of the military that pose a significant threat to global health, especially to civilian health in countries with substantial military presence or underdeveloped health systems. While biological transmission mechanisms are shared by other social groups, selected social transmission mechanisms are unique to the military. As an increasingly interconnected world faces the challenges of COVID-19 and future infectious diseases, the identified features of the military may exacerbate current and similar challenges and impair attempts to implement successful and equitable global public health policies.

## BACKGROUND

With the development of communication and transportation technologies, increase in international trade, and mass population movements, the potential for infectious disease agents to cause global pandemics has increased.^1^ Coronavirus disease 2019 (COVID-19), caused by the Severe Acute Respiratory Syndrome Coronavirus 2 (SARS-CoV2), is a case in point, as this virus has spread faster than the other two recent coronavirus diseases: Severe Acute Respiratory Coronavirus (SARS-CoV) and Middle East Respiratory Syndrome Coronavirus (MERS-CoV). In an increasingly interconnected world, understanding the transmission mechanisms of emerging viruses, as well as vulnerabilities and gaps in current public health measures, is crucial to developing effective and equitable public health policy.

Initial restrictions on the movement of populations contributed to flattening the global disease curve of COVID-19.^2^ Overtime, widespread repurposing of existing drugs have led to important drops in morbidity and mortality,^3,4,5^ a better understanding of the pathophysiology of COVID-19 is helping to stratify and individualize treatment strategies,^6,7^ and vaccine developments are providing hope. However, one key transmission vector has been overlooked by government officials, policymakers, and scientists alike in their policy responses, nationally and globally: the role of the military as a disease vector. Its underreporting notwithstanding, there is well documented evidence, spanning over a century, for the military as a pathogen transmitter.^8^ For example, the so-called Spanish Flu infected around 500 million people, one third of the world’s population at the time, killing at least 50 million - by some counts around 100 million. Despite its name, recent historiography suggests that this pandemic originated not in Spain but in the United States of America (USA), in Camp Funston, Fort Riley, Kansas, with US soldiers carrying it to Europe as they crossed the Atlantic to join allied troops in the First World War.^9^ Another instance of military transmission is the case of sexually transmitted diseases (STDs), which ravaged both military personnel and Korean civilians living close to or within US military camp towns (*kijichon*), between the end of the Korean War and late into the 20^th^ century.^10^

Infectious diseases like STDs are not unique to war zones but inherent to the demographics and lifestyles of the military. As the *Military Times* recently noted, US military towns have among the highest rates of STDs, likely due to the young age of service members.^11^ Military recruits are also at high risk of meningococcal disease,^12^ a life-threatening infection with long-term sequelae, associated with young age, high carriage rates due to crowded living quarters, and global deployment to disease endemic regions.

In sum, numerous historical and ongoing outbreaks of infectious diseases have been documented among military personnel. However, to the best of our knowledge only one systematic review on this topic has been conducted to date, albeit before the onset of COVID-19, drawing from only one database, limited to the time frame 1955 - 2018, and including only 67 articles,^8^ thus the goal of this systematic review: to identify, with no temporal restrictions and drawing from three major repositories of medical literature, circumstances under which military-civilian transmission might occur, shed light on both biological and social transmission mechanisms, and elaborate on the implications of distinct features of the military for global public health policy.

## METHODS

Our study followed the Preferred Reporting Items for Systematic Reviews and Meta-Analysis (PRISMA) guidelines^13^ for conducting reviews in healthcare. The protocol was registered with the International Prospective Register of Systematic Review (PROSPERO) (registration number: CRD42020188699).

### Search strategy

Our overarching research question was: “What are the biosocial mechanisms whereby disease transmission occurs within the military and between military and civilian populations?” On May 13, 2020, we conducted a search in 3 electronic databases (Ovid MEDLINE, Ovid EMBASE, and Web of Science) using combinations of Medical Subject Headings (MeSH) and keyword search terms with no temporal restrictions. Key words included “military”, “army”, “troops”, “navy”, “naval base”, “soldier”, “disease vector”, “disease carrier”, “disease transmission”, “pathogen transmission”, “epidemic”, “outbreak”, “infect”, “civilian” (full search strategy available under “supplementary materials”). We supplemented our database search by scanning the reference lists of included studies. Because one important aim of our study was to understand its implications for COVID-19, we also manually searched the grey literature (Google Scholar) to identify COVID-19 studies in military populations that addressed our research question.

### Selection criteria and screening

We included original research studies if they (1) were peer-reviewed and (2) provided evidence or supporting information for the military as a disease vector, or for military missions as high-risk environments/settings for the spread of infectious diseases, or (3) provided evidence for the spread of disease within the military, or (4) provided evidence for the spread of disease between military and civilian populations. We excluded articles if they (1) were not in English, (2) were reviews, case studies, letters, conference abstracts, editorials, commentaries, or surveillance reports, (3) did not describe/explain features of the military that promoted the spread of infectious disease or (4) did not use human participants. Authors independently screened each study in two separate rounds of study selection, a first consisting of title and abstract screening and a second consisting of full text screening. Disagreements were resolved by consensus.

### Data extraction

We used a pre-formatted Excel worksheet (Microsoft, Redmond, Washington, USA) to extract data from studies meeting inclusion criteria. Extracted data included information such as study characteristics (e.g., study type), data collection methods (e.g., survey), sample size and participant composition (e.g., military vs civilian), associated countries (e.g., country of military origin), disease incidence characteristics (e.g., total cases, proportions among subgroups), and disease transmission characteristics (e.g., biological vs social mechanisms).

### Data analysis

We applied an inductive narrative synthesis approach combining content analysis and thematic analysis to assess, summarize, and appraise findings that addressed our research question.^14^ Upon identifying biological mechanisms of transmission, we grouped studies according to their social mechanisms of transmission by extracting quotes to identify themes and sub-themes. To demonstrate strength of support, we generated frequency distributions of themes and sub-themes.^14^ We assumed trustworthiness based on accepted standards of trustworthiness in qualitative research - credibility, dependability, confirmability, transferability; and authenticity.^15^

## RESULTS

### Included studies

Our search identified 6477 articles. After removing duplicates and non-English records, 3597 articles remained for screening. Upon title and abstract screening, we excluded 2651 articles, which left 946 for full-text review, based on which we excluded 738 articles, thus leaving 208 that met our inclusion criteria. Our grey literature search yielded 2 additional articles on COVID-19 in the military. Our inter-rater reliability for article screening was 82%. Figure 1 summarizes the flow of literature searching and screening.

**FIGURE 1.**
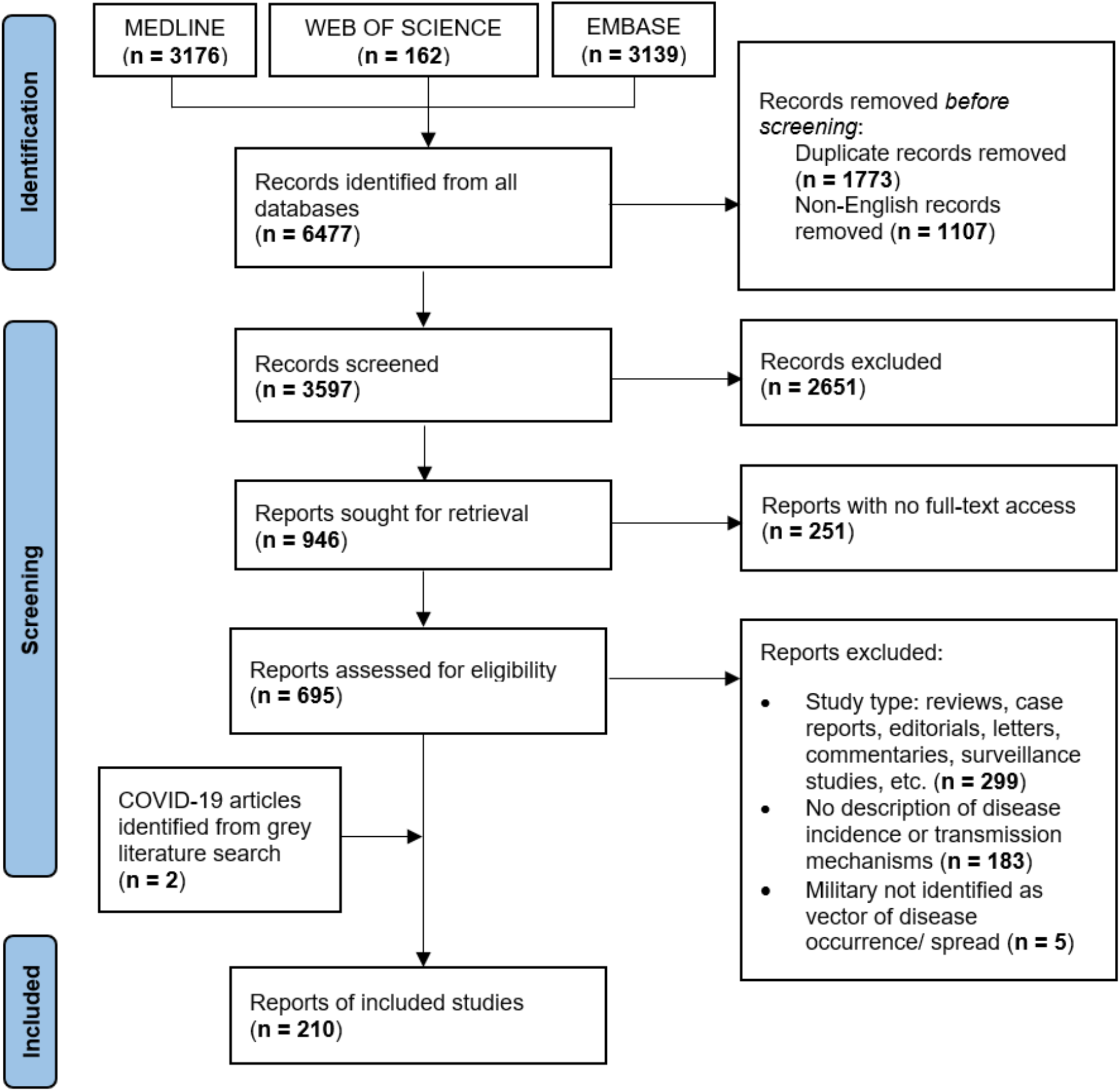
PRISMA Flow chart for study selection

### Study characteristics

Included records were published between 1810 and 2020, with a wide range of data collection periods (1 day - 24 years), sample sizes (48 - 8990 participants), and locations (67 countries), with most studies conducted in North America (Figure 2), specifically the USA (20%; 43/210), and the military most frequently originating in North America (Figure 2), specifically the USA (34%; 72/210). When comparing regions of study location (Figure 2a) with regions of military origin (Figure 2b) or of author affiliation (Figure 2c), findings indicated that many studies took place in Asia or Africa, yet with far fewer author affiliations or military origins in those regions (Table 1 & Table S1 in supplementary materials).

**TABLE 1.** Summary of selected characteristics of the 210 included studies

| **Characteristic** | **No. (%)** |
| --- | --- |
| **Year of Publication** |  |
| 1800 - 1820 | 1 (0.5) |
| 1821 - 1840 | 0 (0) |
| 1841 - 1860 | 1 (0.5) |
| 1861 - 1880 | 0 (0) |
| 1881 - 1900 | 0 (0) |
| 1901 - 1920 | 2 (1) |
| 1921 - 1940 | 1 (0.5) |
| 1941 - 1960 | 7 (3) |
| 1961 - 1980 | 14 (7) |
| 1981 - 2000 | 33 (16) |
| 2001 - 2020 | 151 (72) |
| **Populations** |  |
| Military | 173 (82) |
| Military and civilian | 34 (16) |
| Civilian | 2 (1) |
| Unspecified | 1 (0.5) |
| **Military type** |  |
| Army | 89 (42) |
| Training base | 51 (24) |
| Navy | 30 (14) |
| Air Force | 20 (10) |
| Marine Corps | 19 (9) |
| Hospital | 6 (3) |
| Academic institution | 6 (3) |
| Medical corps | 2 (1) |
| Coast Guard | 0 (0) |
| Unspecified | 35 (17) |
| **Method of data collection** |  |
| Laboratory testing | 156 (74) |
| Questionnaire | 112 (53) |
| Interview | 60 (29) |
| Medical record review | 48 (23) |
| Environmental sampling (food or water sources) | 28 (13) |
| Observations (i.e., physical examination) | 14 (7) |
| Focus group | 2 (1) |
| Participant journal/diary | 1 (0.5) |
| **Type of infectious disease** |  |
| Foodborne / waterborne | 84 (40) |
| Droplets | 73 (35) |
| Sexually transmitted and bloodborne infections | 30 (14) |
| Vector borne | 29 (14) |
| Airborne | 15 (7) |
| Close contact<sup>1</sup> | 11 (5) |
| Unspecified | 2 (0.9) |
| **Disease incidence<sup>2</sup>** |  |
| Confirmed<sup>3</sup> incidence from microbiological testing | 136 (65) |
| Suspected<sup>4</sup> incidence only | 27 (13) |
| Incidence not reported | 47 (22) |
| **Disease Transmission Populations** |  |
| Military to military | 183 (87) |
| Military to civilian | 25 (12) |
| Civilian to military | 25 (12) |
| Vector to military | 15 (7) |
| Civilian to civilian | 3 (1) |
| Vector to civilian | 2 (1) |
| Military to vector to military | 1 (0.5) |
| Unspecified | 6 (3) |

<sup>1</sup>Refers to infections that spread through sustained close contact rather than through casual contact (i.e., cold/flu microbes)
<sup>2</sup>Reported incidence is likely not the true incidence as many authors included only patient participants or did not include data for participants lost to follow-up
<sup>3</sup>Methods of confirmation of disease include:
- a) isolation of pathogen from normally sterile site - b) using a plaque reduction neutralization test - c) using a real-time reverse transcription polymerase chain reaction - d) serologically positive for infection as per specific antibody testing
<sup>4</sup>Refers to symptoms of disease without microbiological testing

**FIGURE 2.**
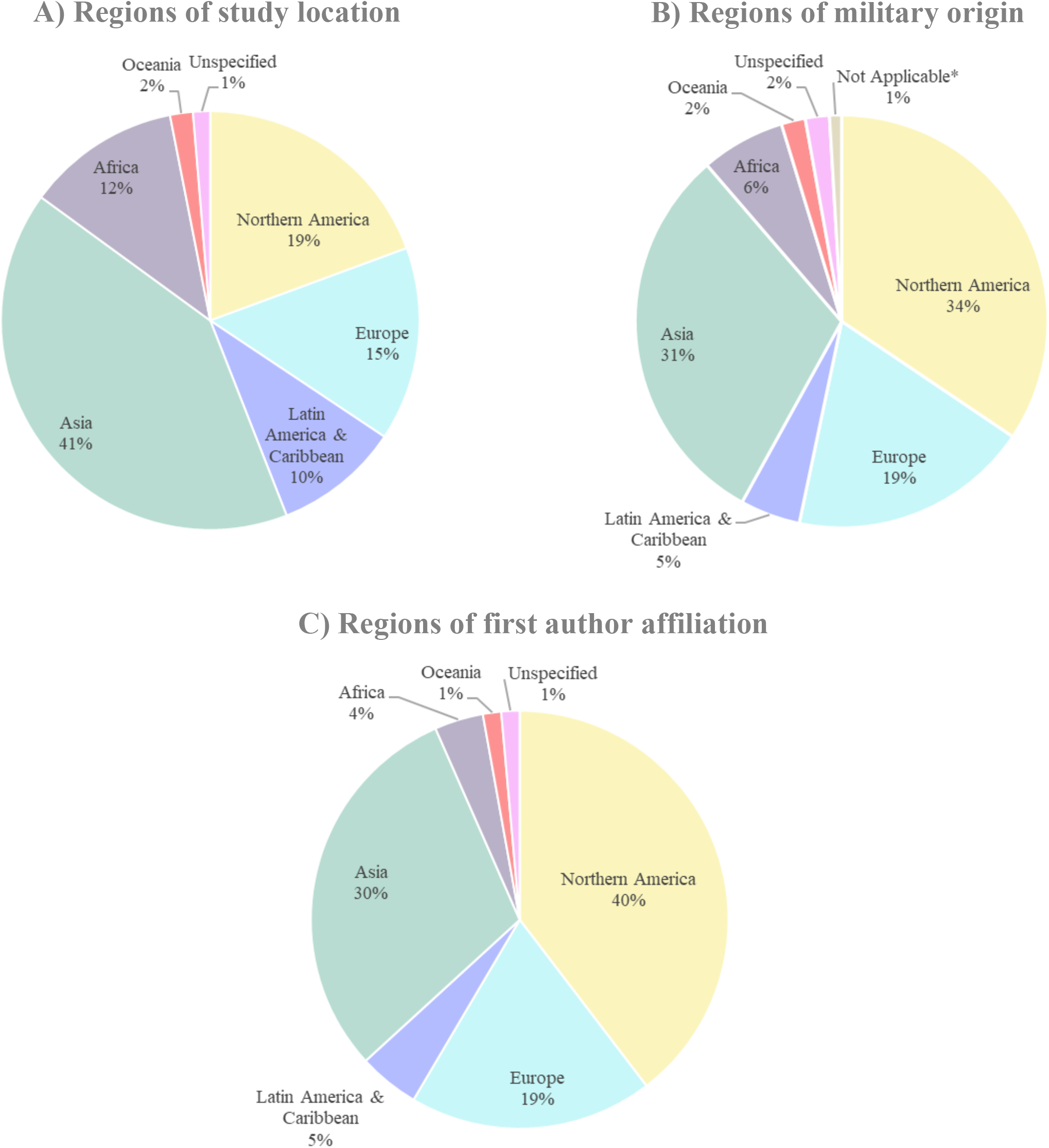
Pie charts showing region-level data pertaining to the studies included in the analysis. We grouped countries into 6 regions: Northern America, Latin America & Caribbean, Asia, Europe, Africa, and Oceania, based on the United Nations geoscheme system. Please see Table S2 in our supplementary materials for the specific list of countries included within each region. Pie chart (A) depicts the percent of studies taking place in each of the 6 regions. Pie chart (B) depicts the percent of studies with military groups originating from each of the 6 regions. Pie chart (C) depicts the percent of studies whose first author is affiliated with each of the 6 regions. Some articles took place in multiple regions, studied military groups originating from multiple regions, and/or the first author had multiple affiliations. \**Not Applicable* refers to articles which did not include the military among their study populations.

Of the 210 articles, only 17% (36/210) studied civilians, of which 33% (12/36) discussed the impact of military outbreaks on civilians, with only 1% (3/210) of studies finding that disease incidence among civilians was lower than in the military. Fifteen additional studies (7%; 15/210) did not study civilians but discussed the impact on civilians of military outbreaks. A majority (67%; 140/210) identified the military branch studied, characterized as Army (42%; 89/210), Navy (14%; 30/210), Marine Corps (9%; 19/210), Air Force (10%; 20/210), or Medical Corps (1%; 2/210), with many including more than one branch. Almost one quarter (24%; 51/210) of studies took place within military training bases. A small minority described outbreaks involving military populations at hospitals (3%; 6/210) and academic institutions (3%; 6/210). Three studies (1%; 3/210) involved only civilians or did not specify population type, discussing the military only peripherally.

Data collection methods varied, with many studies employing more than one (Table S1). Most studies (74%; 156/210) employed laboratory testing to identify outbreak causative agent(s) and determine disease incidence. Of 156 studies employing laboratory testing, the vast majority (87%; 136/156) tested to identify the disease agent and/or provide incidence rates (Table 1). Just over half (53%; 112/210) employed questionnaires to determine participant perspectives or knowledge of various diseases. Many questionnaires also collected data on illness, symptoms, places frequented by participants, and other information directly related to identifying sick personnel, disease transmission, and spread. Less used methods included interviews (29%; 60/210), medical record reviews (23%; 48/210), environmental sampling (13%; 28/210), observations (i.e., physical exam) (7%; 14/210), focus groups (1%; 2/210), and participant journals (<1%; 1/210).

### Biological mechanisms of transmission

The most common biological transmission mechanism identified was contaminated food/water, with 40% (84/210) of studies describing foodborne/waterborne-caused outbreaks. Other mechanisms were droplet-transmitted infections (35%; 73/210), sexually transmitted and bloodborne infections (14%; 30/210), vector-borne infections (14%; 29/210), airborne infections (7%; 15/210), and close contact infections (5%; 11/210). Some articles identified more than one biological transmission mechanism, so frequencies do not add up to the total number of articles.

### Social mechanisms of transmission

One hundred and eighty (86%; 180/210) articles reported on social mechanisms of disease transmission. Our thematic analysis identified twelve such mechanisms that we grouped under three categories: (1) policy (i.e., occupation-specific freedom of movement, vaccination programs), (2) institutional (i.e., contractor mismanagement, food contamination, living conditions, pressure from military leadership, poor infrastructure, poor public health management and services, training conditions, working conditions), and (3) individual (i.e., high-risk behaviours, ignoring public health advice). Because articles with quotes that corresponded to more than one social mechanism were counted as reporting on multiple mechanisms (Table S1), social transmission mechanism frequencies do not add up to the total number of articles reporting on them (Table 2).

**TABLE 2.**
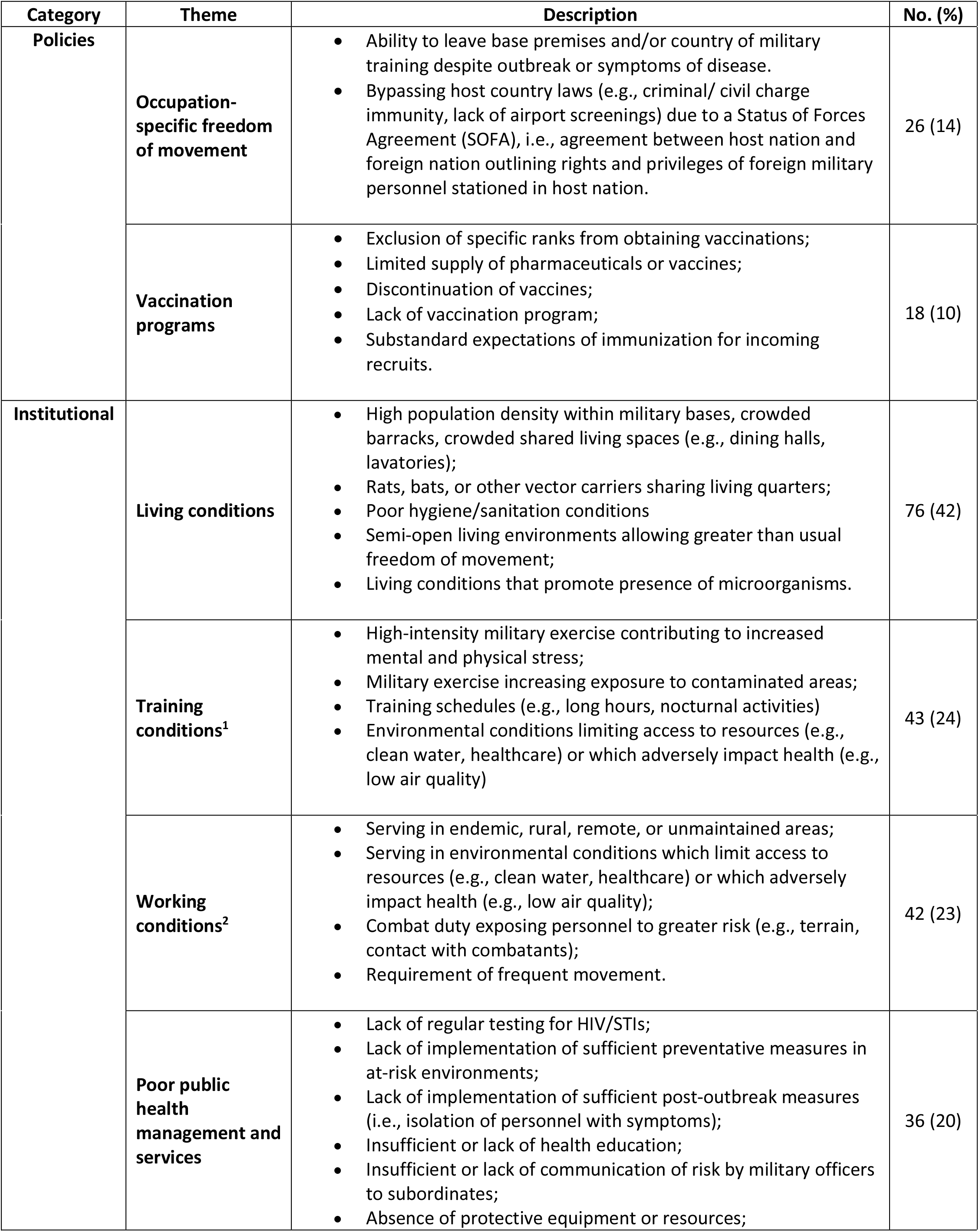
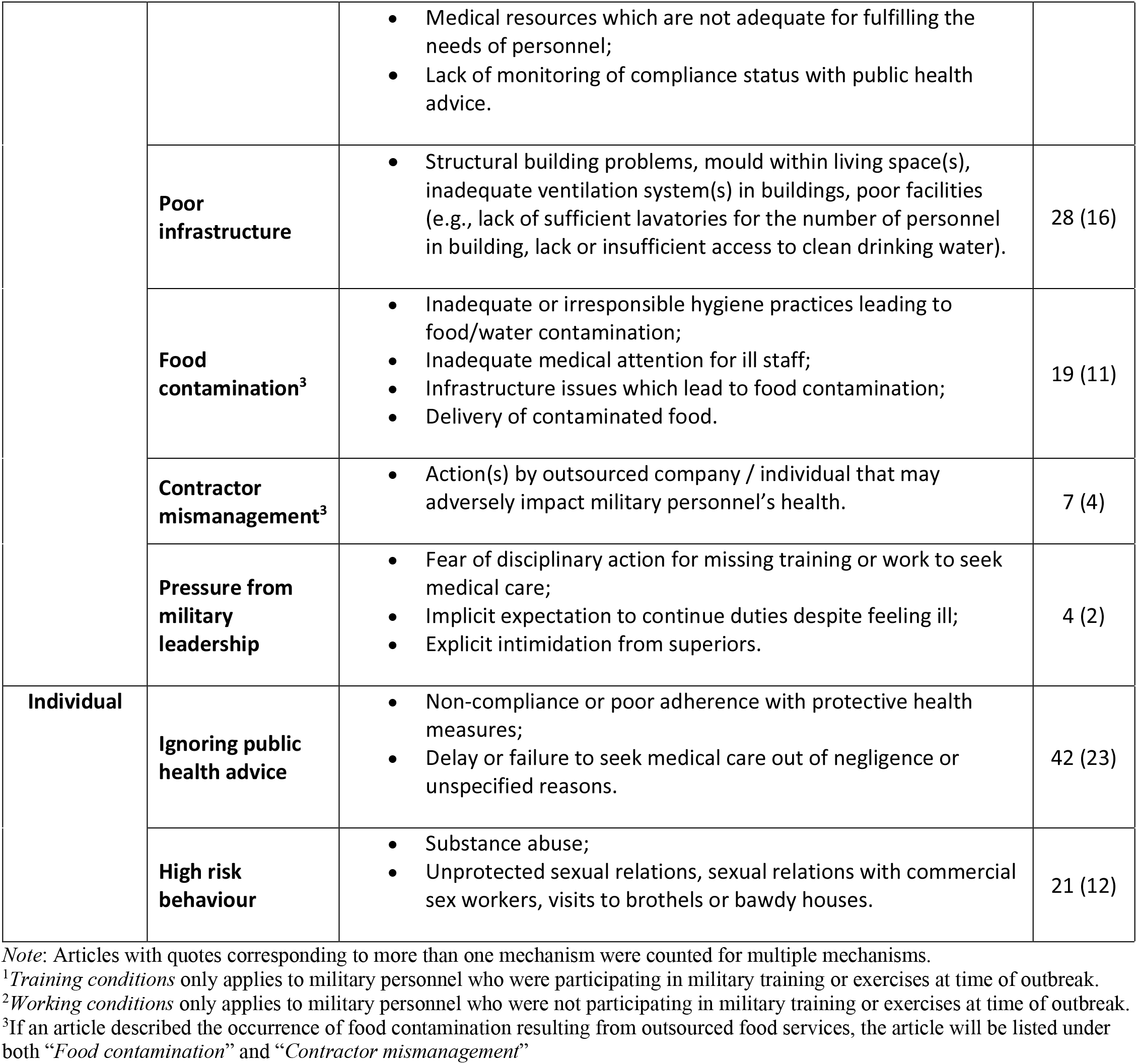
Frequencies of Social Mechanisms of Transmission among included articles

| Category | Theme | Description | No. (%) |
| --- | --- | --- | --- |
| **Policies** | **Occupation-specific freedom of movement** | • Ability to leave base premises and/or country of military training despite outbreak or symptoms of disease.<br>• Bypassing host country laws (e.g., criminal/ civil charge immunity, lack of airport screenings) due to a Status of Forces Agreement (SOFA), i.e., agreement between host nation and foreign nation outlining rights and privileges of foreign military personnel stationed in host nation. | 26 (14) |
|  | **Vaccination programs** | • Exclusion of specific ranks from obtaining vaccinations;<br>• Limited supply of pharmaceuticals or vaccines;<br>• Discontinuation of vaccines;<br>• Lack of vaccination program;<br>• Substandard expectations of immunization for incoming recruits. | 18 (10) |
| **Institutional** | **Living conditions** | • High population density within military bases, crowded barracks, crowded shared living spaces (e.g., dining halls, lavatories);<br>• Rats, bats, or other vector carriers sharing living quarters;<br>• Poor hygiene/sanitation conditions<br>• Semi-open living environments allowing greater than usual freedom of movement;<br>• Living conditions that promote presence of microorganisms. | 76 (42) |
|  | **Training conditions<sup>1</sup>** | • High-intensity military exercise contributing to increased mental and physical stress;<br>• Military exercise increasing exposure to contaminated areas;<br>• Training schedules (e.g., long hours, nocturnal activities)<br>• Environmental conditions limiting access to resources (e.g., clean water, healthcare) or which adversely impact health (e.g., low air quality) | 43 (24) |
|  | **Working conditions<sup>2</sup>** | • Serving in endemic, rural, remote, or unmaintained areas;<br>• Serving in environmental conditions which limit access to resources (e.g., clean water, healthcare) or which adversely impact health (e.g., low air quality);<br>• Combat duty exposing personnel to greater risk (e.g., terrain, contact with combatants);<br>• Requirement of frequent movement. | 42 (23) |
|  | **Poor public health management and services** | • Lack of regular testing for HIV/STIs;<br>• Lack of implementation of sufficient preventative measures in at-risk environments;<br>• Lack of implementation of sufficient post-outbreak measures (i.e., isolation of personnel with symptoms);<br>• Insufficient or lack of health education;<br>• Insufficient or lack of communication of risk by military officers to subordinates;<br>• Absence of protective equipment or resources; | 36 (20) |
|  |  | Medical resources which are not adequate for fulfilling the needs of personnel;<br>Lack of monitoring of compliance status with public health advice. |  |
|  | **Poor infrastructure** | Structural building problems, mould within living space(s), inadequate ventilation system(s) in buildings, poor facilities (e.g., lack of sufficient lavatories for the number of personnel in building, lack or insufficient access to clean drinking water). | 28 (16) |
|  | **Food contamination<sup>3</sup>** | Inadequate or irresponsible hygiene practices leading to food/water contamination;<br>Inadequate medical attention for ill staff;<br>Infrastructure issues which lead to food contamination;<br>Delivery of contaminated food. | 19 (11) |
|  | **Contractor mismanagement<sup>3</sup>** | Action(s) by outsourced company / individual that may adversely impact military personnel's health. | 7 (4) |
|  | **Pressure from military leadership** | Fear of disciplinary action for missing training or work to seek medical care;<br>Implicit expectation to continue duties despite feeling ill;<br>Explicit intimidation from superiors. | 4 (2) |
| **Individual** | **Ignoring public health advice** | Non-compliance or poor adherence with protective health measures;<br>Delay or failure to seek medical care out of negligence or unspecified reasons. | 42 (23) |
|  | **High risk behaviour** | Substance abuse;<br>Unprotected sexual relations, sexual relations with commercial sex workers, visits to brothels or bawdy houses. | 21 (12) |

*Note:* Articles with quotes corresponding to more than one mechanism were counted for multiple mechanisms.
<sup>1</sup>*Training conditions* only applies to military personnel who were participating in military training or exercises at time of outbreak.
<sup>2</sup>*Working conditions* only applies to military personnel who were not participating in military training or exercises at time of outbreak.
<sup>3</sup>If an article described the occurrence of food contamination resulting from outsourced food services, the article will be listed under both "*Food contamination*" and "*Contractor mismanagement*"

#### Policy

##### Occupation-specific freedom of movement

Military personnel are very mobile this mobility is specifically related to the nature of military life and the goals of the military as a social institution: thus military personnel are often required to complete training courses in foreign countries, deployed to foreign bases to fulfill missions, and travel to bases external from their home base.^16^ New recruits regularly enter training bases as others who have completed training leave and personnel are often transferred from one base to another.^17^ Deployed military personnel are not always subjected to similarly comprehensive population health assessments as non-mobile personnel.^18^ Therefore military mobility contributes to spreading infections across populations, with 14% (26/180) of studies reporting on this social mechanism of transmission.

Specifically, studies reported on military personnel assigned to complete multinational exercises,^19-21^ with leave granted upon exercise completion^16,22^ and subsequent travel of suspected cases to other locations, likely spreading disease. Studies also reported on the arrival of returning infected soldiers and infected recruits leading to outbreaks in the study population, or on the transfer of participants during the study resulting in further spread to other locations,^17,23-25^ and on personnel lost to follow-up,^26-28^ so true disease incidence could not be determined. Moreover, a few studies reported on the presence of travelling military personnel in civilian areas (e.g., airports; public transit),^29,30^ providing opportunities for transmission between military and civilian populations. While no study reported on the statistical significance of occupation-related freedom of movement as a factor for disease occurrence, some reported direct temporal associations between the arrival of military personnel from one location and a subsequent outbreak in the location of arrival.^20,31,32^

##### Vaccination programs

Of the 180 studies identifying social mechanisms of transmission, 10% (18/180) reported suboptimal vaccination programs as contributing to disease incidence. Reasons included discontinuation of vaccines by suppliers during, or leading up to, the study period,^31^ low vaccine supply during the study period,^33^ or immunization not required for enrolment.^34^ One study with two groups exposed to an infectious agent reported the outbreak almost entirely in the non-vaccinated group.^35^ Of studies reporting absent or inadequate vaccination programs, 33% (6/18) described military populations with less than 35% of personnel vaccinated against the outbreak-causing disease.

#### Institutional

##### Contractor mismanagement

A few articles (4%; 7/180) reported on ‘contractor mismanagement’, which we defined as any action performed by private contractors that may negatively impact military health. We conceptualized these actions as social mechanisms of disease transmission and included actions such as unhygienic practices by cooks or food handlers contracted by the military,^36,37^ or military-contracted health professionals or food handlers who continued to work despite experiencing symptoms.^38^

##### Food contamination

Although foodborne spread of disease is a biological mechanism of transmission, certain behaviours are required for food contamination. Around one tenth (11%; 19/180) of articles cited food preparation by unfit food handlers (e.g., working despite being symptomatic)^36^ and consumption of food prepared with poorly handled ingredients (e.g., meat left unrefrigerated for long periods)^37,39^ as frequently associated with illness.

##### Living conditions

Living conditions as a probable social mechanism of disease transmission were reported by a large minority of articles (42%; 76/180), including crowded living spaces,^26^ found to be statistically significant for disease acquisition. Other usual but not statistically significant factors included exposure to animals^40^ or insects and unhygienic living quarters.^41^ Articles reporting on living conditions displayed a trend (18%; 14/76), whereby disease was more prevalent among younger, lower ranking and less educated military personnel.^29,35,42^

##### Poor infrastructure

A minority of articles (16%; 28/180) reported on poor infrastructure as contributing to disease spread. The use of contaminated water by military personnel, especially if no other sources were available, was reported as associated with illness,^39,41^ while many other articles reported it as a probable factor.^16,20,43,44^ Additionally, poor facilities, including unsanitary and/or unmaintained latrines,^45^ unchlorinated or inadequately chlorinated water supply,^43^ old and corroded water pipelines,^44^ inadequate ventilation,^32^ poor air quality,^46^ absence of essential appliances (e.g., no refrigeration facility,^45^ no heating appliances^42^) or an insufficient number of facilities^28,29^ were also reported as contributing factors to becoming ill.

##### Poor public health management and services

Although no article reported statistical significance between poor institutional management and/or services and disease incidence, about a fifth (20%; 36/180) reported probable associations related to this theme. Outbreaks also occurred in military bases with poor procedures,^19,21,25^ including lack of testing before leaving and/or after arriving for deployment,^20^ inadequate and/or obsolete supplies for use in military-serving water treatment plants,^39^ delays in placing infected patients in isolation,^38,28^ lack of enforced drug prophylaxis policy^47^, infected personnel allowed to leave the base whilst symptomatic,^16,23,32,48^ and personnel inadequately trained/educated and/or not equipped with the proper equipment/supplies for assigned tasks.^47,49,50^ Additionally, a common contributing factor in training bases was penalizing trainees who missed training, by requiring them, for instance, to restart training, with trainees reporting that they delayed or avoided seeking treatment for this reason despite experiencing symptoms.^40,51^

##### Pressure from military leadership

Very few articles (2%; 4/180) reported on the possible adverse effects of high-pressure often placed on military personnel – especially trainees – to report for duty: these articles reported that military personnel delayed or neglected to seek treatment due to a culture in the military of avoiding to interrupt duties for medical reasons deemed of low to moderate severity.^19^

##### Training conditions

Approximately one fourth of articles (24%; 43/180) reported on training conditions as a social mechanism of transmission. This mechanism is only applicable to the subset of articles taking place in training facilities (24%; 51/210), of which a majority (84%; 43/51) reported on it. Training assignments in remote and/or unmaintained areas (e.g., marshes) and participation in exercises with heavy physical components^24^ were significantly associated with disease transmission. Specifically, travel to endemic areas, nocturnal exercises, low crawl training, sleeping in tents, poor nutrition and/or dehydration, and crowded training bases were identified among possible risk factors for infection.^52,53^

##### Working conditions

In close to a fourth (23%; 42/180) of articles, working conditions were reported as a social mechanism of disease spread. Specifically, crowdedness and being stationed near a stream or river, frequent troop movements, exhaustion, exposure to insects and livestock, and service in disease endemic areas were identified as risk factors.^31,39,40,46^

#### Individual

##### Ignoring public health advice

Military personnel and staff study participants in about one fifth (23%; 42/180) of articles were found to disregard public health advice. A significant association between lack of habitual handwashing prior to cooking by military food preparation staff and increased incidence of disease was reported by one article,^41^ with many others indicating a likely correlation between poor hand hygiene practices and disease.^44^ Similarly, another article found that prophylaxis non-compliance was significantly correlated with disease,^49^ with numerous others identifying prophylaxis non-compliance as a probable factor.^24^ Other behavioural factors likely correlated with infection included lack of use of mosquito nets and approved-grade insect repellent sprays, and failure to wear permethrin-dipped and skin-covering clothing.^19^

##### High-risk behaviour

Demographics and circumstances of military life make military personnel more likely to engage in high-risk behaviours than the general population. Personnel largely consists of young, single men and women who frequently leave their families for long periods for field operations.^54^ Long absences from home tend to increase feelings of loneliness, which correlates with increased likelihood of engaging in risky behaviours (e.g., promiscuous sexual practices, substance abuse).^55^ Although articles did not determine statistical significance between risk behaviours and disease, about one tenth (12%; 21/180) of articles discussing this social mechanism found that disease was more prevalent among individuals who engaged in unprotected sex, heavy smoking, or substance use, compared to those who did not.

### COVID-19 in the military

While our systematic database search did not identify articles with SARS-CoV-2 as the infectious disease agent, our manual grey literature search identified 2 such articles (< 1%; 2/210) meeting our inclusion criteria.^30,56^ These articles documented outbreaks of COVID-19 in the military, one in the United States and one in Niger. Pirnay et al.’s epidemiological analysis of laboratory data suggested that the outbreak originated from a military soldier having direct contact with a local and subsequently infecting other military personnel.^30^ Letizia et al. found that recruits were the most probable source of the disease – particularly two who tested positive for COVID-19 before the outbreak (i.e., on day 0).^56^ These studies also reported on living conditions,^56^ training conditions,^30^ and occupation-specific freedom of movement^30^ as social mechanisms of transmission.

## DISCUSSION

Our systematic review confirms that multiple mechanisms drive disease transmission within military missions, bases, and medical institutions, into civilian populations. Both *biological* mechanisms of transmission - a critical one being contaminated food/water – and *social* mechanisms - such as crowded living, sleeping, and training practices - were common, and are shared by other social groups and institutions. However, selected social mechanisms were unique to the military, such as pressure from military leadership to prioritize military goals over public health safety and occupation-specific freedom of movement. We also found that these social mechanisms have been occurring in military environments as early as 1810.^57^ We posit that they continue into the 21^st^ century despite knowledge of disease containment measures because they are generally accepted as normal and necessary to military goals. We also note that while our findings should not be construed as supporting any specific public health policy, neglecting the role of the military as a pathogen transmitter may have important implications for the wellbeing of communities, for policy formulation, and for global health equity, especially as the military is increasingly assigned tasks overlapping those of humanitarian and medical personnel.^58,59^

Only a few articles studied the impact of disease transmission on civilians (17%; 36/210), despite many reporting information suggesting that civilians were very likely to have been impacted by military outbreaks, such as military personnel granted family leave during the study, dependents living among military personnel, military personnel deployed to bases near civilian populations, and/or military personnel visiting civilian areas (e.g., ports, food vendors, brothels). These findings reveal a trend in the literature whereby studies involving military populations limit their analyses to those populations despite their likely implications on civilians, indicating a gap with potential public health implications. Because studies involving military populations are often conducted by military affiliated researchers, reporting on civilians may be considered beyond the scope of such studies. In fact, the single systematic review on this topic that we identified was not published in a military journal yet was almost entirely devoted to underscoring the threat the circumstances of civilian life and work may pose to service members.^8^ We grant that this is an important point but posit that given the circumstances under which military-to-civilian or civilian-to-military disease spread may occur, these populations are linked and more studies comprehensively investigating transmission among these populations are needed.

Although our included studies describe a myriad of diseases, at the time we conducted our search, the literature still lacked peer-reviewed studies reporting on original research involving COVID-19 outbreaks in the military, with only 2 of 210 on this topic retrieved after an additional search.^30,56^ Nonetheless, reports of military missions acting as disease vectors outside of peer-reviewed literature are quickly accumulating in the current COVID-19 era: for instance, in a period of less than three weeks, more than 40 US Navy warships had at least one sailor test positive.^60^ As recently as March 2021, in South Korea, home of the United States Forces Korea staffed by some 28,000 troops,^61^ a staff member of the military, four servicemembers in quarantine, and one on vacation, tested positive for COVID-19, bringing the total number of infections reported among the military in that country to 658, 31 of whom are undergoing treatment.^62^ In Germany, with over 38,000 stationed US troops,^63^ the commander of a unit in which hundreds of troops contracted COVID-19 soon upon arrival in the country was accused of deploying a leadership style that may have led to his violating quarantine rules.^64^ Meanwhile, Japan was reporting a new cluster of close to 100 COVID-19 cases in military bases in Okinawa, alongside an increase in newly infected civilians in the capital, Tokyo.^65^ These outbreaks within the military may explain at least in part subsequent outbreaks in adjacent populations.

### Limitations

Our review has limitations: we could not calculate disease incidences among study populations because military personnel participants were often transferred, granted leave, or completed training prior to study completion, so participants were lost to follow-up. Therefore, even for studies reporting incidence rates (Table S1), these were likely underreported. Additionally, incidence rates may also be skewed because authors only obtained samples for laboratory testing from very small subsets of populations in a military base or restricted participation to symptomatic subjects. Therefore, the sample size from which incidence was determined was frequently not representative of the actual phenomenon of interest, i.e., disease incidence. Finally, we could not report on the significance of different factors and/or population attributes on disease incidence, since many articles either included participants who had to report for duty at a different military base so left the study setting before completion, or only included symptomatic patients as participants and therefore could not provide a true incidence of disease in the studied population.

## CONCLUSION

Our findings shed light on the role of the military as an important pathogen transmitter, albeit neglected. Unlike biological transmission mechanisms, many social mechanisms that facilitate transmission within the military or between military and civilian populations - pressure from military leadership or occupation-specific freedom of movement – are unique to military life. We have sought to document and call attention to this matter with a view to contributing to the formulation, development, and implementation of more effective and equitable global public health policy.

## Data Availability

All data generated or analysed during this study are available within the article and its supplementary information files.

## Abbreviations

COVID-19: Coronavirus disease 2019
MeSH: Medical Subject Headings
MERS-CoV: Middle East respiratory syndrome coronavirus
PRISMA: Preferred Reporting Items for Systematic Reviews and Meta-Analyses
PROSPERO: International Prospective Register of Systematic Review
SARS-CoV: Severe acute respiratory coronavirus
SARS-CoV2: Severe acute respiratory syndrome coronavirus 2
STD: Sexually transmitted disease
USA: United States of America

## DECLARATIONS

### Ethics approval and consent to participate

Not applicable.

### Consent for publication

Not applicable.

### Authors’ contributions

CC conceptualized and designed the study, registered the review in PROSPERO, conducted the literature review, oversaw all steps of the study, initiated and led the writing of the manuscript, and secured funding for the project. IAD and HF assisted with the development of the study design, conducted searches, screened studies, extracted data from included studies, created tables and figures, drafted the appendices, and assisted with the drafting and revisions of the manuscript. SM extracted data from included studies, assisted with drafting appendices, and reviewed and revised the manuscript. KJN co-originated the study idea, informed the literature review, and revised drafts. All authors approved the final manuscript as submitted.

## Acknowledgements

The first author wishes to thank her students and collaborators, who are a continuing source of inspiration for her work on global health policy and social justice. All authors wish to thank the School of Health Policy and Management for supporting collaborations between faculty members and emerging scholars.

## References

1 Saker L, Lee K, Cannito B, Gilmore A, Campbell-Lendrum, D. Globalization and infectious diseases: a review of the linkages. Geneva, Switzerland: World Health Organization; 2004 https://www.who.int/tdr/publications/documents/seb_topic3.pdf

2 Zhou Y, Xu R, Hu D, Yue Y, Li Q, Xia J. Effects of human mobility restrictions on the spread of COVID-19 in Shenzhen, China: a modelling study using mobile phone data. Lancet Digit Health. 2020;2(8):e417–e424. doi:10.1016/S2589-7500(20)30165-5

3 Bryant A, Lawrie TA, Dowswell T, et al. Ivermectin for prevention and treatment of COVID-19 infection: a systematic review, meta-analysis, and trial sequential analysis to inform clinical guidelines. Am J Ther. 2021;28(4):e434–e460. doi: 10.1097/MJT.0000000000001402.

4 Kory P, Meduri GU, Varon J, Iglesias J, Marik PE. Review of the emerging evidence demonstrating the efficacy of ivermectin in the prophylaxis and treatment of COVID-19. Am J Ther. 2021;28(3):e299–e318. doi:10.1097/MJT.0000000000001377

5 Carvallo H, Roberto H, Psaltis A, Contreras V. Study of the efficacy and safety of topical ivermectin + iota-carrageenan in the prophylaxis against COVID-19 in health personnel. J Biomed Res Clin Invest. 2020;2(1). https://media.marinomed.com/8b/7a/c7/nota-journal-of-biomedical-research-safety-adn-efficacy-iota-carrageenan-and-ivermectin.pdf

6 Marik PE, Iglesias J, Varon J, Kory P. A scoping review of the pathophysiology of COVID-19. Int J Immunopathol Pharmacol. 2021;35:1–16. doi: 10.1177/20587384211048026

7 Risch HA. Early outpatient treatment of symptomatic, high-risk COVID-19 patients that should be ramped up immediately as key to the pandemic crisis. Am J Epidemiol. 2020;189(11):1218–1226. doi:10.1093/aje/kwaa093

8 Zemke JN, Sanchez JL, Pang J, Gray GC. The double-edged sword of military response to societal disruptions: a systematic review of the evidence for military personnel as pathogen transmitters. J Infect Dis. 2019;220(12):1873–1884. doi:10.1093/infdis/jiz400

9 Barry JM. The site of origin of the 1918 influenza pandemic and its public health implications. J Transl Med. 2004;2(1):3. doi:10.1186/1479-5876-2-3

10 Hughes DM, Chon KY, Ellerman DP. Modern-day comfort women: the U.S. military, transnational crime, and the trafficking of women. Violence Against Women. 2007;13(9):901–922. doi:10.1177/1077801207305218

11 Kime P. These military towns have the highest rates of sexually transmitted diseases in the country. Military Times. January 15, 2020. Accessed May 26, 2021. https://www.militarytimes.com/news/your-military/2020/01/15/these-military-towns-have-the-highest-rates-of-sexually-transmitted-diseases-in-the-country/

12 Brundage JF, Ryan MAK, Feighner BH, Erdtmann FJ. Meningococcal disease among United States military service members in relation to routine uses of vaccines with different serogroup-specific components, 1964–1998. Clin Infect Dis. 2002;35(11):1376–1381. doi:10.1086/344273

13 Page MJ, McKenzie JE, Bossuyt PM, et al. The PRISMA 2020 statement: an updated guideline for reporting systematic reviews. BMJ. 2021;372:n71. doi:10.1136/bmj.n71

14 Popay J, Roberts H, Sowden A, et al. Guidance on the conduct of narrative synthesis in systematic reviews: a product from the ESRC methods programme. 2006. doi: 10.13140/2.1.1018.4643

15 Dixon-Woods M, Sutton A, Shaw R, et al. Appraising qualitative research for inclusion in systematic reviews: a quantitative and qualitative comparison of three methods. J Health Serv Res Policy. 2007;12(1):42–47. doi:10.1258/135581907779497486

16 Tong C, Javelle E, Grard G, et al. Tracking rift valley fever: from Mali to Europe and other countries, 2016. Euro Surveill. 2019;24(8):1800213. doi:10.2807/1560-7917.ES.2019.24.8.1800213

17 Cosby MT, Pimentel G, Nevin RL, et al. Outbreak of H3N2 influenza at a US military base in Djibouti during the H1N1 pandemic of 2009. PLoS One. 2013;8(12):e82089. doi:10.1371/journal.pone.0082089

18 Harbertson J, Scott PT, Moore J, et al. Sexually transmitted infections and sexual behaviour of deploying shipboard US military personnel: a cross-sectional analysis. Sex Transm Infect. 2015;91(8):581–588. doi: Journal of Pakistan Association of Dermatologists 10.1136/sextrans-2015-052163

19 Liu W, Kizu JR, Le Grand LR, et al. Localized outbreaks of epidemic polyarthritis among military personnel caused by different sublineages of ross river virus, Northeastern Australia, 2016–2017. Emerg Infect Dis. 2019;25(10):1793–1801. doi:10.3201/eid2510.181610

20 Frerichs RR, Keim PS, Barrais R, Piarroux R. Nepalese origin of cholera epidemic in Haiti. Clin Microbiol Infect. 2012;18(6):E158–E163. doi:10.1111/j.1469-0691.2012.03841.x

21 Ejaz A, Raza N, Din QU, Bux H. Outbreak of cutaneous leishmaniasis in Somniani, Balochistan – implementation of preventive measures for deployed personnel of armed forces. J Pak Assoc of Dermatol. 2008;18:220–225. http://citeseerx.ist.psu.edu/viewdoc/download?doi=10.1.1.452.3871&rep=rep1&type=pdf

22 Harris PNA, Oltvolgyi C, Islam A, et al. An outbreak of scrub typhus in military personnel despite protocols for antibiotic prophylaxis: doxycycline resistance excluded by a quantitative PCR-based susceptibility assay. Microbes Infect. 2016;18(6):406–411. doi:10.1016/j.micinf.2016.03.006

23 Neo FJX, Loh JJP, Ting P, et al. Outbreak of caliciviruses in the Singapore military, 2015. BMC Infect Dis. 2017;17(1):719. doi:10.1186/s12879-017-2821-y

24 Suryam V, Bhatti VK, Kulkarni A, Mahen A, Nair V. Outbreak control of community acquired pneumonia in a large military training institution. Med J Armed Forces India. 2015;71(1):33–37. doi:10.1016/j.mjafi.2014.09.015

25 Schmid D, Kasper S, Kuo HW, et al. Ongoing rubella outbreak in Austria, 2008-2009. Euro Surveill. 2009;14(16):19184. doi:10.2807/ese.14.16.19184-en

26 Yoon CG, Kang DY, Jung J, et al. The infectivity of pulmonary tuberculosis in Korean army units: evidence from outbreak investigations. Tuberc Respir Dis (Seoul). 2019;82(4):298–305. doi:10.4046/trd.2018.0077

27 Dierks J, Servies T, Do T. A study on the leptospirosis outbreak among US Marine trainees in Okinawa, Japan. Mil Med. 2018;183(3-4):e208–e212. doi:10.1093/milmed/usx013

28 Dahanayaka NJ, Kiyohara T, Agampodi SB, et al. Clinical features and transmission pattern of hepatitis A: an experience from a hepatitis A outbreak caused by two cocirculating genotypes in Sri Lanka. Am J Trop Med Hyg. 2016;95(4):908–914. doi:10.4269/ajtmh.16-0221

29 Thornton S, Davies D, Chapman F, et al. Detection of Norwalk-like virus infection aboard two U.S. Navy ships. Mil Med. 2002;167(10):826–830. https://pubmed.ncbi.nlm.nih.gov/12392249/

30 Pirnay J-P, Selhorst P, Cochez C, et al. Study of a SARS-CoV-2 outbreak in a Belgian military education and training center in Maradi, Niger. Viruses. 2020;12(9):949. doi:10.3390/v12090949

31 McNeill KM, Ridgely Benton F, Monteith SC, Tuchscherer MA, Gaydos JC. Epidemic spread of adenovirus type 4-associated acute respiratory disease between U.S. Army installations. Emerg Infect Dis. 2000;6(4):415–419. doi:10.3201/eid0604.000419

32 Lawson R. Observations on the outbreak of yellow fever among the troops at Newcastle, Jamaica, in the latter part of 1856. Br Foreign Med Chir Rev. 1859;24(48):445–480. https://www.ncbi.nlm.nih.gov/pmc/articles/PMC5182490/pdf/brforeignmcrev72734-0165.pdf

33 Halhal B, Glick Y, Galor I, Ran A, Bacon DJ, Glassberg E. Pertussis outbreak among soldiers during basic training: the need for updated protocols. Mil Med. 2017;182(S1):355–359. doi:10.7205/MILMED-D-16-00083

34 Seah SG-K, Lim EA-S, Kok-Yong S, et al. Viral agents responsible for febrile respiratory illnesses among military recruits training in tropical Singapore. J Clin Virol. 2010;47(3):289–292. doi:10.1016/j.jcv.2009.12.011

35 Blouse LE, Kolonel LN, Corrado V. Influenza A/England: an outbreak at a military academy. Am J of Epidemiol. 1974;100(3):216–221. doi:10.1093/oxfordjournals.aje.a112030

36 Watier-Grillot S, Boni M, Tong C, et al. Challenging investigation of a norovirus foodborne disease outbreak during a military deployment in Central African Republic. Food Environ Virol. 2017;9(4):498–501. doi:10.1007/s12560-017-9312-6

37 Kasper MR, Lescano AG, Lucas C, et al. Diarrhea outbreak during U.S. military training in El Salvador. PLoS One. 2012;7(7):e40404. doi:10.1371/journal.pone.0040404

38 Lessa FC, Gould PL, Pascoe N, et al. Health care transmission of a newly emergent adenovirus serotype in health care personnel at a military hospital in Texas, 2007. J Infect Dis. 2009;200(11):1759–1765. doi:10.1086/647987

39 Aho M, Kurki M, Rautelin H, Kosunen TU. Waterborne outbreak of campylobacter enteritis after outdoors infantry drill in Utti, Finland. Epidemiol Infect. 1989;103(1):133–141. doi:10.1017/S0950268800030430

40 Faix DJ, Harrison DJ, Riddle MS, et al. Outbreak of Q fever among US military in Western Iraq, June–July 2005. Clin Infect Dis. 2008;46(7):e65–e68. doi:10.1086/528866

41 Brainard J, D’hondt R, Ali E, et al. Typhoid fever outbreak in the Democratic Republic of Congo: case control and ecological study. PLoS Negl Trop Dis. 2018;12(10):e0006795. doi:10.1371/journal.pntd.0006795

42 Kushwaha AS, Aggarwal SK, Arora MM. Outbreak of meningococcal infection amongst soldiers deployed in operations. Med J Armed Forces India. 2010;66(1):4–8. doi:10.1016/S0377-1237(10)80082-6

43 Tuck JJH, Williams JR, Doyle AL. Gastro enteritis in a military population deployed in West Africa in the UK Ebola response; was the observed lower disease burden due to handwashing? Travel Med and Infect Dis. 2016;14(2):131–136. doi:10.1016/j.tmaid.2015.12.009

44 Singh P, Handa SK, Banerjee A RETD. Epidemiological investigation of an outbreak of viral hepatitis. Med J Armed Forces India. 2006;62(4):332–334. doi:10.1016/S0377-1237(06)80100-0

45 Kuhns DM, Anderson TG. A fly-born bacillary dysentery epidemic in a large military organization. Am J Public Health Nations Health. 1944;34(6):750–755. doi:10.2105/ajph.34.7.750

46 Sanchez JL, Binn LN, Innis BL, et al. Epidemic of adenovirus-induced respiratory illness among US military recruits: epidemiologic and immunologic risk factors in healthy, young adults. J Med Virol. 2001;65(4):710–718. doi:10.1002/jmv.2095

47 Sanchez JL, Bendet I, Max Grogl L, et al. Malaria in Brazilian military personnel deployed to Angola. J Travel Med. 2000;7(5):275–282. doi:10.2310/7060.2000.00077

48 Farrell M, Sebeny P, Klena JD, et al. Influenza risk management: lessons learned from an A(H1N1) pdm09 outbreak investigation in an operational military setting. PLoS One. 2013;8(7):e68639. doi:10.1371/journal.pone.0068639

49 Fernando SD, Booso R, Dharmawardena P, et al. The need for preventive and curative services for malaria when the military is deployed in endemic overseas territories: a case study and lessons learned. Mil Med Res. 2017;4(1):19. doi:10.1186/s40779-017-0128-3

50 Azuogu B, Ogbonnaya L, Alo C. HIV voluntary counseling and testing practices among military personnel and civilian residents in a military cantonment in southeastern Nigeria. HIV AIDS (Auckl). 2011;3:107–116. doi:10.2147/HIV.S23774

51 Hammond-Collins K, Strauss B, Barnes K, et al. Group A streptococcus outbreak in a Canadian Armed Forces training facility. Mil Med. 2019;184(3-4):e197–e204. doi:10.1093/milmed/usy198

52 Brosch L, Tchandja J, Marconi V, et al. Adenovirus serotype 14 pneumonia at a basic military training site in the United States, spring 2007: a case series. Mil Med. 2009;174(12):1295–1299. doi:10.7205/milmed-d-03-0208

53 Vainio A, Lyytikäinen O, Sihvonen R, et al. An outbreak of pneumonia associated with S. pneumoniae at a military training facility in Finland in 2006. APMIS. 2009;117(7):488–491. doi:10.1111/j.1600-0463.2009.02463.x

54 Mgbere O, Monjok E, Abughosh S, Ekong E, Holstad MM, Essien EJ. Modeling covariates of self-perceived and epidemiologic notions of risk for acquiring STIs/HIV among military personnel: a comparative analysis. AIDS Behav. 2013;17(3):1159–1175. doi:10.1007/s10461-011-0126-5

55 Mankayi N. Military men and sexual practices: discourses of ‘othering’ in safer sex in the light of HIV/AIDS. SAHARA J. 2009;6(1):33–41. doi:10.1080/17290376.2009.9724927

56 Letizia AG, Ramos I, Obla A, et al. SARS-CoV-2 transmission among marine recruits during quarantine. N Eng J Med. 2020;383(25):2407–2416. doi:10.1056/NEJMoa2029717

57 Lichtenstein H. Account of the epidemic dysentery which prevailed among the Dutch troops at the Cape of Good Hope, in 1804 and 1805. Edinb Med Surg J. 1810;6(23):296–305. https://www.ncbi.nlm.nih.gov/pmc/articles/PMC5747859/pdf/edinbmedsurgj71835-0036.pdf

58 Herhalt C. These are the five struggling long-term care homes the military has been sent to help. CTV News. April 24, 2020. Accessed April 21, 2021. https://toronto.ctvnews.ca/these-are-the-five-struggling-long-term-care-homes-the-military-has-been-sent-to-help-1.4910112?cache=yes

59 Michaud J, Moss K, Licina D, et al. Militaries and global health: peace, conflict, and disaster response. Lancet. 2019;393(10168):276–286. doi:10.1016/S0140-6736(18)32838-1

60 Starr B. How the coronavirus pandemic has shaken the US military. CNN. April 26, 2020. Accessed April 21, 2021.https://www.cnn.com/2020/04/25/politics/coronavirus-impact-us-military/index.html

61 Harkins G. About 28,000 US troops are stationed in South Korea. Only 28 got COVID-19. Military.com. March 21, 2021. Accessed July 31, 2021. https://www.military.com/daily-news/2021/03/11/about-28000-us-troops-are-stationed-south-korea-only-28-got-covid-19.html

62 All News. USFK apologizes for ‘no mask’ dance parties amid pandemic. Yonhap News Agency. December 9, 2020. Accessed December 20, 2020. https://en.yna.co.kr/view/AEN20201209008300325

63 Knight B. US military in Germany: what you need to know. Deutsche Welle. June 16, 2020. Accessed July 31, 2021. https://www.dw.com/en/us-military-in-germany-what-you-need-to-know/a-49998340.

64 Vandiver J. Fort Hood-based brigade commander under investigation after allegations of toxic leadership, flouting coronavirus rules. Stars and Stripes. March 29, 2021. Accessed March 29, 2021. https://www.stripes.com/fort-hood-based-brigade-commander-under-investigation-after-allegations-of-toxic-leadership-flouting-coronavirus-rules-1.667669

65 Ditzler J. US military on Okinawa tightens coronavirus restrictions as case numbers increase. Stars and Stripes. March 30, 2021. Accessed April 23, 2021. https://www.stripes.com/news/pacific/us-military-on-okinawa-tightens-coronavirus-restrictions-as-case-numbers-increase-1.667774

