## Supplemental Material for "The military as a neglected pathogen transmitter, from the 19th century to COVID-19: A systematic review" (from another version of this preprint: version 1, 2021-10-12)

**Supplementary Materials**

**TABLE S1** **– Search strategy**

Strategy 1

| **Embase Search 1** | | |
| --- | --- | --- |
| **#** | **Searches** | **Results** |
| 1 | military.mp. or army/ | 73405 |
| 2 | ("military base" or "military personnel").mp. | 6704 |
| 3 | (troops or navy or "naval base").mp. | 6830 |
| 4 | soldier/ | 28479 |
| 5 | "disease vector".mp. | 998 |
| 6 | disease carrier/ | 32015 |
| 7 | disease transmission/ | 97408 |
| 8 | "pathogen transmission".mp. | 1920 |
| 9 | epidemic/ or outbreak.mp. | 138482 |
| 10 | 1 or 2 or 3 or 4 | 88635 |
| 11 | 5 or 6 or 7 or 8 or 9 | 248215 |
| 12 | 10 and 11 | 2468 |

Strategy 2

| **Embase Search 2** | | |
| --- | --- | --- |
| **#** | **Searches** | **Results** |
| 1 | military.mp. or army/ | 73405 |
| 2 | ("military base" or "military personnel").mp. | 6704 |
| 3 | (troops or navy or "naval base").mp. | 6830 |
| 4 | soldier/ | 28479 |
| 5 | civilian.mp. | 11563 |
| 6 | (outbreak or epidemic or infect*).mp. | 2869602 |
| 7 | 1 or 2 or 3 or 4 | 88635 |
| 8 | 5 and 6 and 7 | 671 |

**Medline**

Strategy 1

| **MEDLINE Search 1** | | |
| --- | --- | --- |
| **#** | **Searches** | **Results** |
| 1 | military.mp. or Military Personnel/ | 87882 |
| 2 | "military base".mp. | 139 |
| 3 | "naval base".mp. | 67 |
| 4 | troops.mp. | 2245 |
| 5 | navy.mp. | 4253 |
| 6 | disease vector.mp. or Disease Vectors/ | 8762 |
| 7 | Disease Transmission, Infectious/ or Disease Outbreaks/ | 86800 |
| 8 | ("disease transmission" or "disease carrier" or "pathogen transmission").mp. | 37101 |
| 9 | (epidemic or outbreak).mp. | 137563 |
| 10 | 1 or 2 or 3 or 4 or 5 | 90644 |
| 11 | 6 or 7 or 8 or 9 | 213820 |
| 12 | 10 and 11 | 2539 |

Strategy 2

| **MEDLINE Search 2** | | |
| --- | --- | --- |
| **#** | **Searches** | **Results** |
| 1 | military.mp. or Military Personnel/ | 87882 |
| 2 | "military base".mp. | 139 |
| 3 | "naval base".mp. | 67 |
| 4 | troops.mp. | 2245 |
| 5 | navy.mp. | 4253 |
| 6 | civilian*.mp. | 11465 |
| 7 | outbreak.mp. or Disease Outbreaks/ | 103465 |
| 8 | epidemic.mp. or Epidemics/ | 91189 |
| 9 | infect*.mp. | 2209877 |
| 10 | 1 or 2 or 3 or 4 or 5 | 90644 |
| 11 | 7 or 8 or 9 | 2297942 |
| 12 | 6 and 10 and 11 | 637 |

**Web of Science**

| **#** | **Searches** | **Results** |
| --- | --- | --- |
| 1 | TOPIC: (military or troops or navy or "naval base") AND TOPIC: (civilian*) AND TOPIC: (outbreak* or epidemic*) | 177 |

**Refined by**: [excluding] DOCUMENT TYPES: (Proceeding Paper or Meeting Abstract)

**Timespan**: All years. **Indexes**: SCI-EXPANDED, SSCI

**TABLE S2** – List of characteristics for each included record

| **First Author,  Year** | **Study Location** | **First Author**  **Location** | **Military Origin** | **Pop.**  **of Interest** | **Military Type** | **Data Collection Methods** | **Sample Size** | **Infectious Disease** | **Biological Mechanism** | **Social Mechanism** |
| --- | --- | --- | --- | --- | --- | --- | --- | --- | --- | --- |
| Abdulla, 2018^1^ | Iraq | Iraq | Iraq | Military, civilians | Army | Interviews, observations | 234 | Leishmaniasis | Vector | Working conditions |
| Agan, 2013^2^ | US | US | US | Military | Army, Marine Corps, Navy | Chart review | 200 | Warts | Sexually transmitted | None |
| Aho, 1989^3^ | Finland | Finland | Finland | Military | Army | Interviews, laboratory tests | 125 | Campylo-  bacteriosis | Food/water | Poor infrastructure, working conditions, ignoring public health advice, food contamination |
| Aho, 2010^4^ | Finland | Finland | Finland | Military | N/S | Laboratory tests, questionnaires, chart review | 346 | Flu | Droplet | Living conditions, poor public health management and services |
| Ali, 2017^5^ | DRC | Belgium | DRC | Military | N/S | Interviews, laboratory tests | 320 | Typhoid fever | Food/water | Poor infrastructure, living conditions, working conditions, occupation-specific freedom of movement |
| Allen, 1975^6^ | US | US | US | Military | Army | Chart review, interviews | N/S | Hepatitis B | Sexually transmitted | High-risk behaviour, poor public health management and service |
| Almog, 1994^7^ | Israel | Israel | Israel | Military | Academic institution | Laboratory tests | N/S | Meningococcal disease | Close contact | Living conditions |
| Altshuler, 1959^8^ | Germany | US | US | Military, civilians | Army | Laboratory tests, observations, environmental sampling | N/S | Bacterial diarrheal illness | Food/water | Poor public health management and service |
| Ambrose, 2014^9^ | US | US | US | Military | N/S | Chart review, interviews, laboratory tests, environmental sampling | 67 | Legionnaires' disease | Droplet | Poor infrastructure |
| Arnold, 2005^10^ | US | US | US | Military | Academic institution | Questionnaires | 483 | Gastroenteritis | Food/water | None |
| Azuogu, 2011^11^ | Nigeria | Nigeria | Nigeria | Military, civilians | Army | Questionnaires | 350 | AIDS | Sexually transmitted | High-risk behaviour, ignoring public health advice, poor public health management and service |
| Bailey, 2005^12^ | Iraq | UK | UK | Military | Hospital | Chart review, questionnaires, laboratory tests | 500 | Gastroenteritis | Food/water | Living conditions, working conditions, poor public health management and service |
| Bakhireva, 2004^13^ | Ethiopia | US | Ethiopia | Military | Air Force, army, training base, hospital, academic institution | Interviews, questionnaires | 289 | AIDS | Sexually transmitted | Working conditions, high-risk behaviour, poor public health management and service |
| Balicer, 2005^14^ | Israel | Israel | Israel | Military | N/S | Chart review, laboratory tests | 48 | Flu | Droplet | Living conditions, vaccination program |
| Banerjee, 2005^15^ | India | India | India | Military | Training base | Laboratory tests, questionnaires | 190 | Hepatitis E | Food/water | Poor infrastructure |
| Banerjee, 2005^16^ | India | India | India | Military | Training base | Chart review, laboratory tests | 131 | Pneumonia | Droplet | Poor infrastructure, living conditions |
| Banerjee, 2007^17^ | India | India | India | Military | Academic institution | Laboratory tests | 163 | Rubella | Food/water | Living conditions, training conditions, occupation-specific freedom of movement |
| Banerjee, 2007^18^ | India | India | India | Military | Training base | Laboratory tests, chart review | 95 | Typhoid fever | Food/water | Poor infrastructure |
| Bar-Dayan, 1996^19^ | Israel | Israel | Israel | Military, civilians | Air Force | Laboratory tests, questionnaires | 146 | Strep throat and impetigo | Droplet | Ignoring public health advice, food contamination |
| Basiliere, 1968^20^ | US | US | US | Military | Navy, Marine Corps, training base | Chart review | 95 | Pneumonia | Droplet | Living conditions, training conditions |
| Bellanger, 2011^21^ | Ivory Coast | Switzerland | France | Military | N/S | Questionnaires | 43 | Malaria | Vector | Working conditions, ignoring public health advice, poor public health management and service |
| Benenson, 1982^22^ | Panama | US | US | Military | Army | Chart review, questionnaires | N/S | Toxoplasmosis | Food/water | Poor infrastructure, training conditions, food contamination |
| Block, 1999^23^ | Israel | Israel | Israel | Military | Army | Interviews, laboratory tests | 1632 | Meningococcal disease | Close contact | Living conditions |
| Blouse, 1974^24^ | US | US | US | Military | Air Force, Navy | Chart review, interviews | N/S | Flu | Droplet | Living conditions, vaccination program |
| Brainard, 2018^25^ | DRC | UK | Democratic Republic of  Congo | Military | N/S | Interviews, laboratory tests, environmental sampling | 960 | Typhoid fever | Food/water | Poor infrastructure, living conditions, ignoring public health advice |
| Brett-Major, 2012^26^ | US | US | US | Military | Navy, Marine Corps | Laboratory tests, questionnaires | 496 | AIDS | Sexually transmitted | None |
| Brockmann, 2008^27^ | Germany | Germany | Germany | Military | N/S | Laboratory tests, environmental sampling, questionnaires | 625 | Cryptosporidiosis | Food/water | Poor infrastructure, training conditions, food contamination |
| Broderick, 2008^28^ | US | US | US | Military | Marine Corps | Laboratory tests, environmental sampling | 13,114 | Adenovirus illness | Droplet | Living conditions |
| Brosch, 2009^29^ | US | US | US | Military | Air Force (training base) | Chart review, laboratory tests | 57 | Adenovirus illness | Droplet | Living conditions, training conditions |
| Brosh-Nissimov, 2018^30^ | Israel | Israel | Israel | Military | Medical corps | Interviews, laboratory tests | 501 | Tinea corporis | Close contact | Working conditions |
| Bryan, 2002^31^ | Pakistan | US | Pakistan | Military | Academic institution | Laboratory tests, environmental sampling, questionnaires | 109 | Hepatitis E | Food/water | None |
| Campbell, 2004^32^ | US | US | US | Military | Training base | Laboratory tests, questionnaires | 202 | Skin infections, flu-like | Close contact | Living conditions, training conditions |
| Casey, 1996^33^ | Peru | Peru | Peru | Military | Army | Laboratory tests | 88 | Hepatitis B, D | Sexually transmitted | Training conditions |
| Cates, 1975^34^ | Germany | US | US | Military | Army | Interviews | 427 | Hepatitis B | Sexually transmitted | High-risk behaviour |
| Cecil, 1919^35^ | US | US | US | Military | N/S | Laboratory tests | 673 | Pneumonia | Droplet | Vaccination program |
| Celentano, 1998^36^ | Thailand | Thailand | Thailand | Military | Army, Air force | Interviews, laboratory tests | 4086 | AIDS | Sexually transmitted | High-risk behaviour |
| Chen, 2010^37^ | Singapore | Singapore | Singapore | Military, civilians | N/S | Laboratory tests, questionnaires | 2909 | Cold and flu-like diseases | Droplet | Working conditions |
| Clayson, 1998^38^ | Nepal | Thailand | Nepal | Military | Army (training base) | Laboratory tests, chart review | N/S | Hepatitis E | Food/water | Poor infrastructure |
| Cohen, 1987^39^ | Israel | Israel | Israel | Military, civilians | Army (training base) | Chart review, interviews, laboratory tests | 151 | Pharyngitis | Food/water | Ignoring public health advice, food contamination |
| Cohen, 2002^40^ | Israel | Israel | Israel | Military | Army | Laboratory tests | 444 | Gastroenteritis | Food/water | Training conditions |
| Cosby, 2013^41^ | Djibouti | Egypt | US | Military, civilians | Navy | Laboratory tests | 32 | Cold and flu-like diseases | Droplet | Living conditions, working conditions, occupation-specific freedom of movement |
| Coursaget, 1998^42^ | Djibouti | France | France | Military, civilians | N/S | Laboratory tests | 172 | Hepatitis A, E | Food/water | None |
| Courtney, 2017^43^ | South Sudan | US | South Sudan | Military | Army | Interviews, laboratory tests | 1063 | AIDS | Sexually transmitted | High-risk behaviour |
| Cowan, 1987^44^ | Germany | US | US | Military | Army | Chart review, interviews | N/S | Hepatitis B | Sexually transmitted | None |
| Cross, 1992^45^ | Mediterranean ports, Pacific ports | US | US | Military | Navy | Questionnaires | N/S | Respiratory illness | Droplet | Living conditions |
| Cruickshank, 1982^46^ | UK | UK | UK | Military | Training base | Laboratory tests, questionnaires | N/S | Strep throat and impetigo | Droplet | Living conditions, training conditions, ignoring public health advice, poor public health management and service |
| Crum, 2002^47^ | US | US | US | Military | Navy | Interviews, observations, questionnaires, chart review, laboratory tests | 22 | Coccidioidomycosis (valley fever) | Airborne | Training conditions |
| Crum, 2004^48^ | US | US | US | Military | Training base | Laboratory tests, chart review | 3500 | Pneumonia | Droplet | Living conditions |
| Crum, 2005^49^ | US | US | US | Military | Marine Corps (training base) | Laboratory tests, questionnaires | 493 | Pneumonia (Group A Streptococcus) | Droplet | Ignoring public health advice |
| Dahanayaka, 2016^50^ | Sri Lanka | Sri Lanka | Sri Lanka | Military | Army | Interviews | 222 | Hepatitis A | Food/water | Poor infrastructure, working conditions, vaccination program, poor public health management and service, occupation-specific freedom of movement |
| De Santi, 2011^51^ | Chad | France | France | Military | Army, Air force, Medical corps | Laboratory tests, questionnaires | 196 | Cold and flu-like diseases | Droplet | Living conditions |
| De Santi, 2016^52^ | French Guiana | France | France | Military | Army | Laboratory tests, environmental sampling | 272 | Malaria | Vector | Working conditions, ignoring public health advice |
| Demoncheaux, 2012^53^ | Senegal | Senegal | France | Military | Army | Laboratory tests, environmental sampling, questionnaires | 149 | Gastroenteritis | Food/water | Contractor mismanagement |
| Dierks, 2018^54^ | Japan | US | US | Military | Marine Corps | Interviews, questionnaires | 239 | Leptospirosis | Food/water | Training conditions, poor public health management and service, occupation-specific freedom of movement |
| Dongliu, 2016^55^ | China | China | China | Military | Training base | Laboratory tests | 1600 | Adenovirus illness | Droplet | Training conditions, poor public health management and service |
| Dudley, 1927^56^ | New Zealand | N/S | New Zealand, UK | Military | Navy | Observations | N/S | Flu | Droplet | None |
| Earhart, 2001^57^ | US | US | US | Military | Navy | Interviews, laboratory tests, questionnaires | 949 | Flu | Droplet | Living conditions, working conditions, vaccination program |
| Ejaz, 2008^58^ | Pakistan | Pakistan | Pakistan | Military | Army | Questionnaires | 108 | Leishmaniasis | Vector | Ignoring public health advice, poor public health management and service, occupation-specific freedom of movement |
| Elazar, 2015^59^ | Israel | Israel | Israel | Military | N/S | Chart review | 1,192 | N/S | N/S | None |
| Essien, 2006^60^ | Nigeria | US | Nigeria | Military | N/S | Questionnaires | 2,214 | AIDS | Sexually transmitted | Working conditions |
| Faix, 2008^61^ | Iraq | US | US | Military | Marine Corps | Chart review, questionnaires, interviews, laboratory tests, environmental sampling | 38 | Q fever | Airborne | Living conditions, working conditions, ignoring public health advice, pressure from military leadership, poor public health management and service |
| Farrell, 2013^62^ | Kuwait | US | US | Military | Army | Laboratory tests, questionnaires | 217 | Flu | Droplet | Working conditions, occupation-specific freedom of movement |
| Feikin, 1999^63^ | N/S | US | N/S | Military | Training base | Chart review, laboratory tests, questionnaires | 586 | Pneumonia, adenovirus illness | Droplet | Living conditions |
| Fernando, 2017^64^ | Sri Lanka | Sri Lanka | Sri Lanka | Military | Air Force | Focus groups, questionnaires | 120 | Malaria | Vector | Working conditions, ignoring public health advice, poor public health management and service |
| Frerichs, 2012^65^ | Haiti | US | Nepal | Military | N/S | Interviews, laboratory tests, observations | N/S | Cholera | Food/water | Poor infrastructure, working conditions, poor public health management and service, occupation-specific freedom of movement |
| Furesz, 2004^66^ | Hungary | Hungary | Hungary | Military | Army | Laboratory tests | 2880 | Gastritis, peptic ulcer disease | Food/water | Living conditions |
| Gallimore, 2005^67^ | Persian Gulf | UK | UK | Military, civilians | Navy | Laboratory tests, questionnaires | N/S | Gastroenteritis, diarrhoea | Food/water | None |
| Gambel, 1999^68^ | Haiti | US | United Nations | Military, civilians | Army | Laboratory tests, questionnaires | 249 | Dengue fever | Vector | Ignoring public health advice, poor public health management and service |
| Gavan, 1970^69^ | France | US | US | Military, civilians | Army | Interviews | 52 | Hepatitis (type unspecified) | Food/water | None |
| Ghose, 2006^70^ | India | India | India | Military, civilians | Army | Laboratory tests, interviews | 91 | Malaria | Vector | Poor infrastructure, working conditions, poor public health management and service |
| Gonzaga, 2011^71^ | Peru | Peru | US | Military | Navy | Laboratory tests, questionnaires | 130 | Gastroenteritis | Food/water | Living conditions, working conditions |
| Gray, 1991^72^ | US | US | US | Military | Marine Corps | Laboratory tests, questionnaires | 736 | Skin infections, flu-like infections | Close contact | Training conditions, ignoring public health advice |
| Gray, 2000^73^ | US | US | US | Military | Army, Navy, Marine Corps | Laboratory tests, chart review | 3413 | Adenovirus illness | Droplet | Vaccination program |
| Gremillion, 1978^74^ | US | US | US | Military | Air Force (training base) | Chart review, interviews, laboratory tests | N/S | Rubella | Droplet | Living conditions |
| Grotto, 2004^75^ | Israel | Israel | Israel | Military | Army (training base) | Interviews, environmental sampling, laboratory tests | 84 | Gastroenteritis | Food/water | Ignoring public health advice, food contamination |
| Hadad, 2006^76^ | Israel | Israel | Israel | Military | Army | Interviews, laboratory tests | 27 | Leptospirosis | Food/water | Training conditions |
| Halhal, 2017^77^ | Israel | Israel | Israel | Military | Training base | Questionnaires | 1596 | Pertussis (whooping cough) | Droplet | Living conditions, vaccination program |
| Hammond-Collins, 201^78^ | Canada | Canada | Canada | Military | Training base | Laboratory tests, interviews | 710 | Strep throat and impetigo | Droplet | Ignoring public health advice, pressure from military leadership, poor public health management and service |
| Harbertson, 2013^79^ | Rwanda | US | Rwanda | Military | Army | Laboratory tests, questionnaires | 1307 | AIDS | Sexually transmitted | High-risk behaviour |
| Harbertson, 2015^80^ | US | US | US | Military | Navy, Marine Corps | Questionnaires | 2453 | STIs | Sexually transmitted | High-risk behaviour, occupation-specific freedom of movement |
| Harbertson, 2019^81^ | US | US | US | Military | Navy, Marine Corps | Questionnaires | 2314 | N/S | Sexually transmitted | None |
| Harris, 2016^82^ | Australia | Australia | Australia | Military | Army | Laboratory tests | 124 | Scrub typhus | Vector | Training conditions, ignoring public health advice, poor public health management and service, occupation-specific freedom of movement |
| Hart, 1974^83^ | Papua New Guinea | Papua New Guinea | Papua New Guinea | Military, civilians | Army | Interviews | 530 | Venereal disease | Sexually transmitted | High-risk behaviour, occupation-specific freedom of movement |
| Hennessy, 2004^84^ | Wales | Wales | Wales | Military | Army (training base) | Laboratory tests, environmental sampling, questionnaires | 105 | Campylobacteriosis | Food/water | Training conditions, ignoring public health advice |
| Hernandez, 1966^85^ | US | US | US | Military, civilians | Academic institution | Interviews | 15 | Hepatitis (type unspecified) | Food/water | Food contamination |
| Hierholzer, 1974^86^ | Spain | US | Spain | Military | N/S | Laboratory tests | 25 | Flu, adenovirus illness | Droplet | None |
| Ho, 2015^87^ | Singapore | Singapore | Singapore | Military | N/S | Environmental sampling, laboratory tests, questionnaires | 5700 | Gastroenteritis | Food/water | Living conditions |
| Hoshino, 2018^88^ | Japan | Japan | N/A | N/S | N/S | Laboratory tests | 29 | Hepatitis C | Sexually transmitted | None |
| Huerta, 2000^89^ | Israel | Israel | Israel | Military, civilians | Army | Laboratory tests, interviews, environmental sampling, questionnaires | 175 | Bacterial diarrheal illness | Food/water | Poor infrastructure |
| Hwang, 2013^90^ | South Korea | South Korea | South Korea | Military | Training base | Laboratory tests | 712 | Cold and flu-like diseases | Droplet | Training conditions |
| Izurieta, 2009^91^ | Brazil | US | N/S | Military | N/S | Interviews, laboratory tests | 341 | Yellow fever | Vector | Training conditions |
| Jadhav, 2007^92^ | India | India | India | Military, civilians | Army | Interviews, environmental sampling, laboratory tests, questionnaires | 254 | Salmonellosis, typhoid fever | Food/water | Food contamination, contractor mismanagement |
| Jeger, 2011^93^ | Switzerland | Switzerland | Switzerland | Military | Army (training base) | Chart review, laboratory tests | 750 | Flu | Droplet | Living conditions, vaccination program, ignoring public health advice |
| Jelastopulu, 2006^94^ | Greece | Greece | Greece | Military | Air Force | Interviews, environmental sampling, questionnaires | 166 | Skin infections, flu-like infections | Close contact | Food contamination, contractor mismanagement |
| Jones, 2006^95^ | Peru | US | Peru | Military | Navy | Laboratory tests | 74 | Giardiasis, cyclosporiasis, shigellosis, campylobacteriosis, bacterial diarrheal illness | Food/water | Poor infrastructure, ignoring public health advice, food contamination |
| Kasper, 2012^96^ | El Salvador | Peru | US | Military | Army | Interviews, laboratory tests, questionnaires | 241 | Shigellosis, cryptosporidiosis, cyclosporiasis | Food/water | Food contamination, contractor mismanagement |
| Kennedy, 1912^97^ | India | N/S | N/S | Military | Army | Observations, laboratory tests | N/S | Dengue fever | Vector | None |
| Khaokham, 2013^98^ | US | US | US | Military | Navy | Chart review, questionnaires, laboratory tests | 489 | Flu | Droplet | Living conditions, working conditions, ignoring public health advice |
| Kim, 2013^99^ | South Korea | South Korea | South Korea | Military | N/S | Questionnaires | 395 | Flu | Droplet | Vaccination program |
| Kitsiripornchai, 1998^100^ | Thailand | Thailand, US | Thailand | Military | Army | Questionnaires | 5474 | AIDS | Sexually transmitted | High-risk behaviour |
| Klement, 2003^101^ | Israel | Israel | Israel | Military | Army | Interviews, laboratory tests, questionnaires | 107 | Pertussis (whooping cough) | Droplet | Living conditions, vaccination program, poor public health management and service, occupation-specific freedom of movement |
| Klement, 2006^102^ | Israel | Israel | Israel | Military | Army (training base) | Laboratory tests, questionnaires | 91 | Pneumonia | Droplet | Living conditions |
| Kocik, 2014^103^ | Poland | Poland | Poland | Military | Army | Laboratory tests, questionnaires | 648 | Cold and flu-like diseases | Droplet | Working conditions |
| Kolavic-Gray, 2002^104^ | US | US | US | Military | Army (training base) | Interviews, laboratory tests, questionnaires | 678 | Adenovirus illness | Droplet | Training conditions |
| Konstantinou, 2007^105^ | Cyprus | Greece | Greece | Military | Army | Laboratory tests | 581 | Rift Valley fever | Vector | Living conditions, training conditions |
| Kotwal, 2005^106^ | Afghanistan, Iraq | US | US | Military | Army | Chart review, questionnaires | 725 | Malaria | Vector | Ignoring public health advice |
| Kuhns, 1944^107^ | US | US | US | Military, civilians | Army | Laboratory tests, questionnaires | 2085 | Shigellosis | Food/water | Poor infrastructure, living conditions, food contamination |
| Kunwar, 2013^108^ | India | India | India | Military | Army | Interviews, laboratory tests, observations, environmental sampling | 53 | Salmonellosis, typhoid fever | Food/water | Food contamination |
| Kunwar, 2015^109^ | India | India | India | Military | Training base | Laboratory tests, interviews | 58 | Pneumonia | Droplet | Living conditions, training conditions, poor public health management and service |
| Kushwaha, 2010^110^ | India | India | India | Military | Army (training base) | Laboratory tests, chart review | 17 | Meningococcal disease | Close contact | Poor infrastructure, living conditions |
| Lagler, 2014^111^ | Austria | Austria | Austria | Military, civilians | N/S | Focus groups, laboratory tests | 997 | Hepatitis E | Food/water | None |
| LaMar, 2003^112^ | US | US | US | Military | Navy, Marine Corps | Laboratory tests | 3338 | Tuberculosis | Droplet | Living conditions, ignoring public health advice |
| Laurel, 2001^113^ | US | US | US | Military | Air force | Laboratory tests, questionnaires | 1048 | Flu | Droplet | None |
| Lawson, 1859^114^ | Jamaica | Jamaica | Jamaica | Military | N/S | Observations | N/S | Yellow fever | Vector | Poor infrastructure, living conditions, ignoring public health advice, poor public health management and service, occupation-specific freedom of movement |
| Lee, 2001^115^ | South Korea | South Korea | South Korea | Military, civilians | N/S | Chart review,  questionnaires | N/S | Malaria | Vector | None |
| Lee, 2010^116^ | Singapore | Singapore | Singapore | Military | N/S | Laboratory tests, questionnaires | 237 | Flu | Droplet | Living conditions, occupation-specific freedom of movement |
| Leece, 1959^117^ | N/S | UK | UK | Military | Army (training base) | Laboratory tests | N/S | Shigellosis | Food/water | None |
| Lesens, 2007^118^ | France | France | France | Military | Army | Interviews, chart review, questionnaires | 273 | Skin infections, flu-like infections | Close contact | Poor infrastructure, living conditions |
| Lessa, 2009^119^ | US | US | US | Civilians | Hospital | Laboratory tests, questionnaires | 218 | Adenovirus illness | Droplet | Poor public health management and service, contractor mismanagement |
| Letizia, 2020^120^ | US | US | US | Military | Marine Corps | Laboratory tests, questionnaires | 1848 | Respiratory infection | Airborne | Living conditions |
| Levy, 2015^121^ | Thailand | Thailand | Thailand | Military | Army (training base) | Laboratory tests | 340 | Cold and flu-like diseases | Droplet | Living conditions, training conditions, high-risk behaviour, vaccination program |
| Lewis, 2010^122^ | UK | UK | UK | Military, civilians | Hospital | Laboratory tests | 5 | Wound infection, bacteremia, pneumonia, urinary tract infection | Airborne | Poor public health management and service |
| Li, 1958^123^ | China | N/S | China | Military | Academic institution | Observations | 472 | Flu | Droplet | Ignoring public health advice, food contamination |
| Liborio, 2015^124^ | Brazil | Brazil | Brazil | Military | N/S | Chart review | 77 | N/S | N/S | Training conditions |
| Lichtenstein, 1810^125^ | South Africa | N/S | Europe | Military | N/S | Observations | N/S | Dysentery | N/S | Training conditions |
| Lindholm, 2017^126^ | Mexico, Caribbean, Central America, South America | US | US | Military, civilians | N/S | Participant journal/diary,  questionnaires | 277 | Chikungunya, Dengue fever | Vector | Working conditions, occupation-specific freedom of movement |
| Liu, 2019^127^ | Australia | Australia | Australia | Military | Army | Questionnaires | 38 | Ross river fever | Vector | Training conditions, ignoring public health advice, pressure from military leadership, poor public health management and service, occupation-specific freedom of movement |
| Lopes-Joao, 2015^128^ | Portugal | Portugal | Portugal | Military | Army | Questionnaires | 160 | N/S | N/S | Training conditions |
| Lurchachaiwong, 2020^129^ | Thailand | Thailand | US | Military | N/S | Laboratory tests, questionnaires | 48 | Campylobacteriosis, bacterial diarrheal illness, salmonellosis, typhoid fever | Food/water | None |
| Macdonald, 2016^130^ | Norway | Norway | Norway | Military, civilians | N/S | Interviews, environmental sampling, questionnaires | 102 | Yersiniosis | Food/water | Living conditions, food contamination |
| Mankayi, 2009^131^ | South Africa | South Africa | South Africa | Military | Academic institution | Interviews | 14 | AIDS | Sexually transmitted | Working conditions, high-risk behaviour |
| Marshall, 2013^132^ | US | US | US | Military | Army | Questionnaires, interviews | 2,259 | AIDS | Sexually transmitted | High-risk behaviour |
| McKee, 1998^133^ | US | US | US | Military, civilians | Army | Chart review | N/S | Syphilis | Sexually transmitted | High-risk behaviour |
| McNeill, 2000^134^ | US | US | US | Military | Army (training base) | Laboratory tests, questionnaires | 327 | Adenovirus illness | Droplet | Working conditions, vaccination program, occupation-specific freedom of movement |
| Mendez, 2019^135^ | Colombia | Colombia | Colombia | Military | Army | Questionnaires | 295 | Changas disease | Vector | Working conditions |
| Mgbere, 2013^136^ | Nigeria | Nigeria | Nigeria | Military | Training base | Questionnaires | 346 | AIDS | Sexually transmitted | Living conditions, working conditions, high-risk behaviour |
| Michel, 2005^137^ | Ivory Coast | France | France | Military, civilians | Army (training base) | Interviews, laboratory tests, environmental sampling, questionnaires | 98 | Salmonellosis, typhoid fever | Food/water | None |
| Mielke, 1989^138^ | Finland | Finland | Sweden | Military, civilians | N/S | Chart review | N/S | N/S | N/S | None |
| Mitchell, 1946^139^ | US | US | US | Military | Army, Air Force | N/S | N/S | Strep throat, impetigo | Droplet | Working conditions |
| Mubayi, 2018^140^ | Spain, Colombia | US | N/S | Military, civilians | N/S | Chart review | N/S | Leishmaniasis | Vector | Working conditions |
| Mukhopadhya, 2006^141^ | India | India | India | Military | Air Force | Questionnaires | 46 | AIDS | Sexually transmitted | Working conditions, high-risk behaviour, poor public health management and service, occupation-specific freedom of movement |
| Mustafa, 2009^142^ | India | India | India | Military, civilians | Army | Environmental sampling,  questionnaires, laboratory tests | 126 | Skin infections, flu-like infections | Food/water | Food contamination |
| Myint, 2007^143^ | East Timor, Burundi, Iraq, Afghanistan | Thailand | Thailand | Military | Army | Laboratory tests | 1971 | Hepatitis E | Food/water | None |
| Neela, 2019^144^ | Malaysia | Malaysia | Malaysia | Military | Hospital | Interviews, laboratory tests | 12 | Leptospirosis | Food/water | Training conditions, ignoring public health advice, poor public health management and service |
| Neo, 2017^145^ | Singapore | Singapore | Singapore | Military | N/S | Observations, environmental sampling, laboratory tests, questionnaires | 150 | Gastroenteritis | Food/water | Poor public health management and service, occupation-specific freedom of movement |
| Nivesvivat,  2016^146^ | Thailand | Thailand | Thailand | Military | Training base | Environmental sampling, interviews, questionnaires, laboratory tests | 212 | Skin infections, flu-like infections | Close contact | Training conditions |
| Nowosiwsky, 1967^147^ | Vietnam | US | US | Military | Army | Observations, laboratory tests | N/S | Malaria | Vector | Working conditions |
| Nwokoji, 2004^148^ | Nigeria | Nigeria | Nigeria | Military | Navy | Focus groups, interviews, questionnaires | 480 | AIDS | Sexually transmitted | Working conditions, high-risk behaviour, poor public health management and service |
| Ochiai, 2005^149^ | Japan | Japan | Japan | Military | N/S | Interviews, questionnaires | 281 | Food poisoning | Food/water | Ignoring public health advice, food contamination |
| Okeke, 2012^150^ | Nigeria | Nigeria | Nigeria | Military | Army | Questionnaires | 299 | AIDS | Sexually transmitted | High-risk behaviour, ignoring public health advice, poor public health management and service |
| Ore, 2015^151^ | Peru | Peru | Peru | Military | Army (training base) | Interviews, observations, laboratory tests | 52 | Leishmaniasis | Vector | Living conditions, training conditions, ignoring public health advice, poor public health management and service |
| Oristo, 2016^152^ | Finland | Finland | Finland | Military | Army | Laboratory tests, questionnaires | 109 | Gastroenteritis, adenovirus illness | Food/water, droplet | Living conditions |
| Pang, 2015^153^ | Singapore | Singapore | Singapore | Military | Training base | Laboratory tests, questionnaires | 8990 | Cold and flu-like diseases | Droplet | Living conditions, training conditions |
| Papaevangelou, 1969^154^ | Greece | Greece | Greece | Military | Navy | Laboratory tests | 416 | Rubella | Droplet | Training conditions |
| Paul, 1958^155^ | South Korea | US | US, South Korea | Military | Army | Chart review, interviews | 122 | Hemorrhagic fever | Vector | Work conditions |
| Peczenik, 1956^156^ | Austria | US | US | Military | Hospital | Laboratory tests, questionnaires | 83 | N/S | Food/water | Working conditions |
| Petersen, 1989^157^ | US | US | US | Military | Army | Laboratory tests, chart review, questionnaires | 86 | Ehrlichiosis | Vector | Training conditions, ignoring public health advice |
| Pirnay,  2020^158^ | Niger | Belgium | Belgium | Military | Training base | Laboratory tests, chart review | 70 | Respiratory infection | Airborne | Training conditions, occupation-specific freedom of movement |
| Pon, 1990^159^ | South Korea | US | US | Military | Marine Corps | Laboratory tests, questionnaires | 2,053 | Hantavirus  pulmonary syndrome | Airborne | Living conditions, training conditions |
| Prier, 1987^160^ | US | US | US | Military | Army | Laboratory tests, questionnaires | 879 | Hepatitis B | Sexually transmitted | High-risk behaviour |
| Qu, 2010^161^ | China | China | China | Military | N/S | Laboratory tests, questionnaires | 2088 | Skin infections, flu-like infections | Close contact | Living conditions |
| Riddle, 2006^162^ | Persian Gulf | US | US | Military | Navy | Chart review | 44 | Gastroenteritis | Food/water | Living conditions |
| Royal, 2013^163^ | Iraq, Qatar | US | US | Military | Marines | Laboratory tests, questionnaires | 932 | Q fever | Airborne | Living conditions, working conditions |
| Russell, 2003^164^ | Peru | Peru | Peru | Military | Navy | Laboratory tests, questionnaires | 92 | Leptospirosis | Food/water | Training conditions, |
| Ryan, 2002^165^ | US | US | US | Military | Navy (training base) | Laboratory tests, questionnaires | 26799 | Adenovirus illness | Droplet | Vaccination program |
| Sanchez, 2000^166^ | Angola | US | Brazil | Military | Army | Chart review, laboratory tests, questionnaires | 439 | Malaria | Vector | Ignoring public health advice, poor public health management and service |
| Sanchez, 2001^167^ | US | US | US | Military | Army (training base) | Laboratory tests, environmental sampling, questionnaires | 255 | Adenovirus illness | Droplet | Poor infrastructure, living conditions |
| Schmid, 2009^168^ | Austria | Austria | Austria | Military | Army | Interviews | 143 | Rubella | Droplet | Living conditions, poor public health management and services, occupation-specific freedom of movement |
| Seah, 2010^169^ | Singapore | Singapore | Singapore | Military | Training base | Laboratory tests, questionnaires | 1354 | Flu | Droplet | Vaccination program |
| Sebeny, 2012^170^ | Egypt | US | US | Military | Army | Laboratory tests, questionnaires | 1,697 | Flu, gastroenteritis | Droplet, food/water | Living conditions |
| Sena, 2000^171^ | US | US | US | Military, civilians | Army | Chart review | N/S | Chlamydia, gonorrhea | Sexually transmitted | None |
| Sharma, 2019^172^ | India | India | India | Military | Training base | Chart review, environmental sampling, questionnaires | 340 | Hepatitis E | Food/water | Food contamination, contractor mismanagement |
| Sharp, 1995^173^ | Mediterranean Sea | Italy | US | Military | Navy | Laboratory tests, questionnaires | 2767 | Gastroenteritis | Food/water | Poor infrastructure, living conditions, working conditions |
| Singh, 1998^174^ | India | India | India | Military | Army | Interviews, laboratory tests | 103 | Salmonellosis, typhoid fever | Food/water | Poor infrastructure, living conditions, ignoring public health advice, food contamination |
| Singh, 2006^175^ | India | India | India | Military | Training base | Laboratory tests, environmental sampling, questionnaires | 36 | Hepatitis E | Food/water | Poor infrastructure, ignoring public health advice |
| Steinberg, 1969^176^ | US | US | US | Military | Marine Corps | Laboratory tests | N/S | Pneumonia | Droplet | Living conditions, training conditions |
| Struewing, 1990^177^ | US | US | US | Military | Navy (training base) | Interviews,  chart review, questionnaires | 1000 | N/S | Airborne | Living conditions, working conditions |
| Sundnes, 1993^178^ | Ethiopia | Ethiopia | Ethiopia | Military | Army | Laboratory tests | N/S | Louse-borne relapsing fever | Vector | None |
| Supramaniam, 1980^179^ | Malaysia | Malaysia | Malaysia | Military | Air force | Laboratory tests | 86 | Rubella | Droplet | None |
| Suryam, 2015^180^ | India | India | India | Military | Training base | Interviews, observations, laboratory tests | 54 | Pneumonia | Droplet | Living conditions, training conditions, occupation-specific freedom of movement |
| Suwannahitatorn, 2014^181^ | Thailand | Thailand | Thailand | Military | Army | Interviews, observations, questionnaires | 249 | Dermatitis | Close contact | Living conditions, training conditions |
| Szwarcwald, 2005^182^ | Brazil | Brazil | Brazil | Military | Army | Questionnaires | 30970 | AIDS | Sexually transmitted | High-risk behaviour |
| Taamasri, 2002^183^ | Thailand | Thailand | Thailand | Military | Army | Laboratory tests, questionnaires | 616 | Blastocystis  infection | Food/water | Poor infrastructure |
| Tam, 2018^184^ | Thailand | Singapore | Thailand | Military | Army | Laboratory tests | 818 | Cold and flu-like diseases | Droplet | Living conditions, vaccination program |
| Tansel, 2003^185^ | Turkey | Turkey | Turkey | Military | Army | Interviews, laboratory tests, environmental sampling | 168 | Salmonellosis, typhoid fever | Food/water | None |
| Tarabbo, 2011^186^ | Italy | Italy | Italy | Military | Navy | Laboratory tests | 216 | Flu | Droplet | Living conditions, working conditions |
| Tate, 2009^187^ | US | US | US | Military | Air Force (training base) | Chart review, interviews, laboratory tests | 173 | Adenovirus illness | Droplet | Training conditions, ignoring public health advice, pressure from military leadership, poor public health management and service |
| Theamboonlers, 2009^188^ | Thailand | Thailand | Thailand | Military | Navy | Laboratory tests, chart review | 134 | Hepatitis A | Food/water | Living conditions, training conditions, food contamination, occupation-specific freedom of movement |
| Thornton, 2002^189^ | US | US | US | Military | Navy, Marine Corps | Laboratory tests, environmental sampling, questionnaires | 81 | Gastroenteritis | Food/water | Poor infrastructure, living conditions, working conditions, occupation-specific freedom of movement |
| Thornton, 2005^190^ | Iraq | US | US | Military | Marine Corps | Laboratory tests | 129 | Gastroenteritis, shigellosis, campylobacteriosis | Food/water | Poor infrastructure, living conditions |
| Todkill, 2018^191^ | Sierre Leone | UK | UK | Military | N/S | Interviews, laboratory tests | 53 | Shigellosis | Food/water | None |
| Tong, 2019^192^ | France | France | France | Military | Army | Questionnaires | 187 | Rift valley fever | Food/water | Poor infrastructure, living conditions, working conditions, poor public health management and service, occupation-specific freedom of movement |
| Torres-Slimming, 2006^193^ | Peru | Peru | Peru | Military | Navy (training base) | Laboratory tests, questionnaires | 274 | Cyclosporiasis | Food/water | Living conditions. training conditions |
| Tran, 2018^194^ | Malawi | US | Malawi | Military | Army, Navy, Air force | Questionnaires | 944 | AIDS | Sexually transmitted | High-risk behaviour |
| Tuck, 2016^195^ | Sierre Leone, Afghanistan | UK | UK | Military | Navy | Questionnaires | 156 | Gastroenteritis | Food/water | Poor infrastructure, living conditions |
| Vainio, 2009^196^ | Finland | Finland | Finland | Military | Training base | Interviews, laboratory tests | 43 | Pneumonia | Droplet | Living conditions, training conditions |
| Vickery, 2008^197^ | Qatar, Kuwait, Turkey | US | US | Military | Army,  Air force, Marine Corps, Navy | Questionnaires | 3446 | Leishmaniasis | Vector | Ignoring public health advice |
| Wadl, 2009^198^ | Germany | Germany | Germany | Military, civilians | N/S | Questionnaires, laboratory tests, environmental sampling | 178 | Gastroenteritis | Food/water | None |
| Wallace, 1996^199^ | Somalia | US | US | Military | Marine Corps | Interviews, questionnaires, laboratory tests, chart review | 499 | Malaria | Vector | Living conditions, working conditions, ignoring public health advice |
| Walton, 1968^200^ | Panama | US | US | Military | Army (training base) | Chart review | 713 | Leishmaniasis | Vector | None |
| Warner, 1991^201^ | US | US | US | Military, civilians | Air Force | Questionnaires, laboratory tests, chart review, interviews, environmental sampling | 2098 | Gastroenteritis | Food/water | Ignoring public health advice, food contamination |
| Watier-Grillot, 2017^202^ | France | France | Central African Republic | Military | Army | Questionnaires | 244 | Gastroenteritis | Food/water | Food contamination, contractor mismanagement |
| Wentz, 2018^203^ | UK | US | UK | Military | Army (training base) | Questionnaires, chart review | 651 | STIs | Sexually transmitted | None |
| Witkop, 2010^204^ | US | US | US | Military | Air Force | Questionnaires, laboratory tests | 1376 | Cold and flu-like diseases | Droplet | Living conditions |
| Yagupsky, 2016^205^ | Israel | Israel | Israel | Civilians | N/S | Laboratory tests | 51 | Osteomyelitis | Sexually transmitted | Living conditions |
| Yap, 2012^206^ | Singapore | Singapore | Singapore | Military, civilians | N/S | Questionnaires, laboratory tests, environmental sampling | 223 | Gastroenteritis | Food/water | Living conditions |
| Yoon,  2019^207^ | South Korea | South Korea | South Korea | Military | Army | Laboratory tests | 953 | Tuberculosis | Droplet | Living conditions, working conditions, vaccination program, poor public health management and service, occupation-specific freedom of movement |
| Yu, 2013^208^ | China | China | China | Military | Training base | Questionnaires, laboratory tests | 176 | Adenovirus illness | Droplet | Living conditions, vaccination program |
| Ziebold, 2003^209^ | Germany | Germany | Germany | Military | Navy | Questionnaires, laboratory tests, chart review | 281 | Rubella | Droplet | Living conditions, working conditions |
| Zinderman, 2004^210^ | US | US | US | Military | Training base | Chart review, interviews, laboratory tests | 1109 | Skin infections,  flu-like | Close contact | Living conditions, training conditions |

*Abbreviations*: AIDS = acquired immunodeficiency syndrome; DRC = Democratic Republic of the Congo; STI = sexually transmitted diseases;

UK = United Kingdom; US = United States; N/S = Not specified

**Table S3: List of specific countries included within each region as per Figure 2**

| **Region** | **List of included countries** |
| --- | --- |
| North America | Canada, United States |
| Latin America & Caribbean | Brazil, Colombia, El Salvador, French Guinea, Haiti, Jamaica, Panama, Peru |
| Asia | Afghanistan, Arabian Gulf, Cyprus, China, East Timor, Egypt, India, Iraq, Israel, Japan, Kuwait, Malaysia, Nepal, Pakistan, Qatar, Sri Lanka, Singapore, South Korea, Thailand, Turkey, Vietnam |
| Europe | Austria, Finland, France, Germany, Greece, Hungary, Italy, Norway, Poland, Portugal, Spain, Switzerland, United Kingdom, Wales |
| Africa | Burundi, Central African Republic, Chad, Democratic Republic of Congo, Dijibouti, Ethiopia, Ivory Coast, Mali, Malawi, Niger, Nigeria, Rwanda, Senegal, Sierre Lionne, Somalia, South Africa, South Sudan |
| Oceania | Australia, New Zealand, Papa New Guinea |

*Note*: List of countries for each region only include countries referenced in one or more of the articles included in this analysis; therefore, these lists are not comprehensive.

References

1. Abdulla QB, Shabila NP, Al-Hadithi TS. An An outbreak of cutaneous leishmaniasis in Erbil governorate of Iraqi Kurdistan Region in 2015. *J Infect Dev Ctries*. 2018;12(08):600-607. doi:10.3855/jidc.10306

2. Agan BK, Macalino GE, Nsouli-Maktabi H, et al. Human papillomavirus seroprevalence among men entering military service and seroincidence after ten years of service. *MSMR*. 2013;20(2):21-24. https://www.ncbi.nlm.nih.gov/pmc/articles/PMC4519827/

3. Aho M, Kurki M, Rautelin H, Kosunen TU. Waterborne outbreak of campylobacter enteritis after outdoors infantry drill in Utti, Finland. *Epidemiol Infect*. 1989;103(1):133-141. doi:10.1017/S0950268800030430

4. Aho M, Lyytikaïnen O, Nyholm JE, et al. Outbreak of 2009 pandemic influenza A(H1N1) in a Finnish garrison - a serological survey. *Euro Surveill*. 2010;15(45). doi:10.2807/ese.15.45.19709-en

5. Ali E, Bergh RVD, D’hondt R, et al. Localised transmission hotspots of a typhoid fever outbreak in the Democratic Republic of Congo. *Pan Afr Med J*. 2017;28. doi:10.11604/pamj.2017.28.179.10208

6. Allen AM, Irwin GR, Karwacki JJ, Warren DC, Levine RS. Epidemic hepatitis B: a sustained outbreak in a large military population. *Am J Epidemiol*. 1975;102(6):545-552. doi:10.1093/oxfordjournals.aje.a112192

7. Almog R, Block C, Gdalevich M, Lev B, Wiener M, Ashkenazi S. First recorded outbreaks of meningococcal disease in the Israel Defence Force: three clusters due to serogroup C and the emergence of resistance to rifampicin. *Infection*. 1994;22(2):69-71. doi:10.1007/BF01739006

8. Altshuler LN, Hernandez DJ. An epidemiological study of an outbreak of water-borne dysentery. *Am J Public Health Nations Health*. 1959;49(1):82-93. doi:10.2105/AJPH.49.1.82

9. Ambrose J, Hampton LM, Fleming-Dutra KE, et al. Large outbreak of Legionnaires’ disease and Pontiac fever at a military base. *Epidemiol Infect*. 2014;142(11):2336-2346. doi:10.1017/S0950268813003440

10. Arnold K, Drenzek C, Salter M, Arduino M, Noble-Wong J. Outbreak of cutaneous bacillus cereus infections among cadets in a university military program - Georgia, August 2004. *MMWR.* 2005;54(48):1233-1235. https://www.jstor.org/stable/23315842

11. Azuogu B, Ogbonnaya L, Alo C. HIV voluntary counseling and testing practices among military personnel and civilian residents in a military cantonment in southeastern Nigeria. *HIV AIDS (Auckl)*. 2011;3:107-116. doi:10.2147/HIV.S23774

12. Bailey MS, Boos CJ, Vautier G, et al. Gastroenteritis outbreak in British troops, Iraq. *Emerg Infect Dis*. 2005;11(10):1625-1628. doi:10.3201/eid1110.050298

13. Bakhireva LN, Abebe Y, Brodine SK, Kraft HS, Shaffer RA, Boyer CB. Human immunodeficiency virus/acquired immunodeficiency syndrome knowledge and risk factors in Ethiopian military personnel. *Mil Med*. 2004;169(3):221-226. doi:10.7205/MILMED.169.3.221

14. Balicer RD, Huerta M, Levy Y, Davidovitch N, Grotto I. Influenza outbreak control in confined settings. *Emerg Infect Dis*. 2005;11(4):579-583. doi:10.3201/eid1104.040845

15. Banerjee A, Kalghatgi AT, Saiprasad GS, et al. Outbreak of pneumococcal pneumonia among military recruits. *Med J Armed Forces India*. 2005;61(1):16-21. doi:10.1016/S0377-1237(05)80111-X

16. Banerjee A, Kalghatgi AT, Singh P, Nagendra A, Singh Z, Handa SK. Epidemiological investigation of an outbreak of enteric fever. *Med J Armed Forces India*. 2007;63(4):322-324. doi:10.1016/S0377-1237(07)80005-0

17. Banerjee A, Sahni AK, Gupta RM, Grewal VS, Singh Z. Outbreak of rubella among cadets in an academy. *Med J Armed Forces India*. 2007;63(2):141-143. doi:10.1016/S0377-1237(07)80058-X

18. Banerjee A, Sahni AK, Rajiva, Nagendra A, Saiprasad GS. Outbreak of viral hepatitis E in a regimental training centre. *Med J Armed Forces India*. 2005;61(4):326-329. doi:10.1016/S0377-1237(05)80055-3

19. Bar-Dayan Y, Bar-Dayan Y, Klainbaum Y, Shemer J. Food-borne outbreak of streptococcal pharyngitis in an Israeli Airforce base. *Scand J Infect Dis*. 1996;28(6):563-566. doi:10.3109/00365549609037961

20. Basiliere JL, Bistrong HW, Spence WF. Streptococcal pneumonia. Recent outbreaks in military recruit populations. *Am J Med*. 1968;44(4):580-589. doi:10.1016/0002-9343(68)90058-2

21. Bellanger A-P, Faucher J-F, Robedat P, Schmitt A, Millon L, Hoen B. Malaria outbreak in French troops returning from Côte d’Ivoire. *Scand J Infect Dis*. 2011;43(3):230-233. doi:10.3109/00365548.2010.538857

22. Benenson MW, Takafuji ET, Lemon SM, Greenup RL, Sulzer AJ. Oocyst-transmitted toxoplasmosis associated with ingestion of contaminated water. *N Engl J Med*. 1982;307(11):666-669. doi:10.1056/NEJM198209093071107

23. Block C, Gdalevich M, Buber R, Ashkenazi I, Ashkenazi S, Keller N. Factors associated with pharyngeal carriage of Neisseria meningitidis among Israel Defense Force personnel at the end of their compulsory service. *Epidemiol Infect*. 1999;122(1):51-57. doi:10.1017/S0950268898001769

24. Blouse LE, Kolonel LN, Corrado V. Influenza A/England: an outbreak at a military academy. *Am J of Epidemiol*. 1974;100(3):216-221. doi:10.1093/oxfordjournals.aje.a112030

25. Brainard J, D’hondt R, Ali E, et al. Typhoid fever outbreak in the Democratic Republic of Congo: case control and ecological study. *PLoS Negl Trop Dis*. 2018;12(10):e0006795. doi:10.1371/journal.pntd.0006795

26. Brett-Major DM, Hakre S, Naito NA, et al. Epidemiology of contemporary seroincident HIV infection in the Navy and Marine Corps. *Mil Med*. 2012;177(11):1328-1334. doi:10.7205/MILMED-D-12-00299

27. Brockmann SO, Dreweck C, Wagner-Wiening C, et al. Serological and epidemiological analysis of an outbreak of gastroenteritis among military recruits in Germany caused by Cryptosporidium parvum. *Infection*. 2008;36(5):450-457. doi:10.1007/s15010-008-7317-7

28. Broderick MP, Hansen CJ, Russell KL. Exploration of the effectiveness of social distancing on respiratory pathogen transmission implicates environmental contributions. *J Infect Dis*. 2008;198(10):1420-1426. doi:10.1086/592711

29. Brosch L, Tchandja J, Marconi V, et al. Adenovirus serotype 14 pneumonia at a basic military training site in the United States, spring 2007: a case series. *Mil Med*. 2009;174(12):1295-1299. doi:10.7205/milmed-d-03-0208

30. Brosh-Nissimov T, Ben-Ami R, Astman N, Malin A, Baruch Y, Galor I. An outbreak of Microsporum canis infection at a military base associated with stray cat exposure and person-to-person transmission. *Mycoses*. 2018;61(7):472-476. doi:10.1111/myc.12771

31. Bryan JP, Iqbal M, Tsarev S, et al. Epidemic of hepatitis E in a military unit in Abbotrabad, Pakistan. *Am J Trop Med Hyg*. 2002;67(6):662-668. doi:10.4269/ajtmh.2002.67.662

32. Campbell KM, Vaughn AF, Russell KL, et al. Risk factors for community-associated methicillin-resistant Staphylococcus aureus infections in an outbreak of disease among military trainees in San Diego, California, in 2002. *J Clin Microbiol*. 2004;42(9):4050-4053. doi:10.1128/JCM.42.9.4050-4053.2004

33. Casey JL, Niro GA, Engle RE, et al. Hepatitis B virus (HBV)/hepatitis D virus (HDV) coinfection in outbreaks of acute hepatitis in the Peruvian Amazon basin: the roles of HDV genotype III and HBV genotype F. *J Infect Dis*. 1996;174(5):920-926. doi:10.1093/infdis/174.5.920

34. Cates W Jr, Warren JW. Hepatitis B in Nuremberg, Germany. Epidemiology of a drug-associated epidemic. Among US Army soldiers. *JAMA*. 1975;234(9):930-934. doi:10.1001/jama.234.9.930

35. Cecil RL, Vaughan HF. Results of prophylactic vaccination against pneumonia at Camp Wheeler. *J Exp Med*. 1919;29(5):457-483. doi:10.1084/jem.29.5.457

36. Celentano DD, Nelson KE, Lyles CM, et al. Decreasing incidence of HIV and sexually transmitted diseases in young Thai men: evidence for success of the HIV/AIDS control and prevention program. *AIDS.* 1998;12(5):F29-F36. doi:10.1097/00002030-199805000-00004

37. Chen MI, Lee VJ, Lim WY, et al. 2009 influenza A(H1N1) seroconversion rates and risk factors among distinct adult cohorts in Singapore. *JAMA*. 2010;303(14):1383-1391. doi:10.1001/jama.2010.404

38. Clayson ET, Vaughn DW, Innis BL, Shrestha MP, Pandey R, Malla DB. Association of hepatitis E virus with an outbreak of hepatitis at a military training camp in Nepal. *J Med Virol*. 1998;54(3):178-182. doi:10.1002/(sici)1096-9071(199803)54:3<178::aid-jmv6>3.0.co;2-2

39. Cohen D, Ferne M, Rouach T, Bergner-Rabinowitz S. Food-borne outbreak of group G streptococcal sore throat in an Israeli military base. *Epidemiol Infect*. 1987;99(2):249-255. doi:10.1017/S0950268800067716

40. Cohen D, Monroe SS, Haim M, et al. Norwalk virus gastroenteritis among Israeli soldiers: lack of evidence for flyborne transmission. *Infection*. 2002;30(1):3-6. doi:10.1007/s15010-001-1163-1

41. Cosby MT, Pimentel G, Nevin RL, et al. Outbreak of H3N2 influenza at a US military base in Djibouti during the H1N1 pandemic of 2009. *PLoS One*. 2013;8(12):e82089. doi:10.1371/journal.pone.0082089

42. Coursaget P, Buisson Y, Enogat N, et al. Outbreak of enterically-transmitted hepatitis due to hepatitis A and hepatitis E viruses. *J Hepatol*. 1998;28(5):745-750. doi:10.1016/S0168-8278(98)80222-5

43. Courtney LP, Goco N, Woja J, et al. HIV prevalence and behavioral risk factors in the Sudan People’s Liberation Army: data from South Sudan. *PLoS One*. 2017;12(11):e0187689. doi:10.1371/journal.pone.0187689

44. Cowan DN, Prier RE. Descriptive epidemiology of an outbreak of hepatitis B in the U.S. Army, Europe. *Mil Med*. 1987;152(7):345-347. doi:10.1093/milmed/152.7.345

45. Cross ER, Hermansen LA, Pugh WM, White MR, Hayes C, Hyams KC. Upper respiratory disease in deployed U.S. Navy shipboard personnel. *Mil Med*. 1992;157(12):649-651. doi:10.1093/milmed/157.12.649

46. Cruickshank JG, Lightfoot NF, Sugars KH, et al. A large outbreak of streptococcal pyoderma in a military training establishment. *J Hyg (Lond)*. 1982;89(1):9-21. doi:10.1017/S0022172400070492

47. Crum N, Lamb C, Utz G, Amundson D, Wallace M. Coccidioidomycosis outbreak among United States Navy SEALs training in a Coccidioides immitis-endemic Area—Coalinga, California. *J Infect Dis*. 2002;186(6):865-868. doi:10.1086/342409

48. Crum NF, Barrozo CP, Chapman FA, Ryan MAK, Russell KL. An outbreak of conjunctivitis due to a novel unencapsulated Streptococcus pneumoniae among military trainees. *Clin Infect Dis.* 2004;39(8):1148-1154. doi: 10.1086/424522

49. Crum NF, Russell KL, Kaplan EL, et al. Pneumonia outbreak associated with group a Streptococcus species at a military training facility. *Clin Infect Dis*. 2005;40(4):511-518. doi:10.1086/427502

50. Dahanayaka NJ, Kiyohara T, Agampodi SB, et al. Clinical features and transmission pattern of hepatitis A: an experience from a hepatitis A outbreak caused by two cocirculating genotypes in Sri Lanka. *Am J Trop Med Hyg*. 2016;95(4):908-914. doi:10.4269/ajtmh.16-0221

51. de Santi VP, Nicand E, Lagathu G, et al. Incidence, etiology, and determinants associated with acute diarrhea among French forces deployed to Chad. *J Travel Med*. 2011;18(2):115-120. doi:10.1111/j.1708-8305.2010.00490.x

52. de Santi VP, Girod R, Mura M, et al. Epidemiological and entomological studies of a malaria outbreak among French armed forces deployed at illegal gold mining sites reveal new aspects of the disease’s transmission in French Guiana. *Malar J*. 2016;15(1):35. doi:10.1186/s12936-016-1088-x

53. Demoncheaux J-P, Michel R, Mazenot C, et al. A large outbreak of scombroid fish poisoning associated with eating yellowfin tuna (Thunnus albacares) at a military mass catering in Dakar, Senegal. *Epidemiol Infect*. 2012;140(6):1008-1012. doi:10.1017/S0950268811001701

54. Dierks J, Servies T, Do T. A study on the leptospirosis outbreak among US Marine trainees in Okinawa, Japan. *Mil Med*. 2018;183(3-4):e208-e212. doi:10.1093/milmed/usx013

55. Dongliu Y, Guoliang Y, Haocheng X, Shuaijia Q, Li B, Yanglei J. Outbreak of acute febrile respiratory illness caused by human adenovirus B P14H11F14 in a military training camp in Shandong China. *Arch Virol*. 2016;161(9):2481-2489. doi:10.1007/s00705-016-2949-x

56. Dudley SF. An analysis of an influenza epidemic in the New Zealand division of the Royal Navy. *J Hyg (Lond)*. 1927;26(2):132-151. doi:10.1017/S0022172400009001

57. Earhart KC, Beadle C, Miller LK, et al. Outbreak of influenza in highly vaccinated crew of U.S. Navy ship. *Emerg Infect Dis*. 2001;7(3):463-365. doi:10.3201/eid0703.010320

58. Ejaz A, Raza N, Din QU, Bux H. Outbreak of cutaneous leishmaniasis in Somniani, Balochistan – implementation of preventive measures for deployed personnel of armed forces. *J Pak Assoc of Dermatol*. 2008;18:220-225. http://citeseerx.ist.psu.edu/viewdoc/download?doi=10.1.1.452.3871&rep=rep1&type=pdf

59. Elazar S, Zelikovich Y, Levine H, Tzurel-Ferber A, Galor I, Hartal M. Secular trends in 1,192 diarrheal outbreaks in the Israel Defence Forces between 1988–2011. *Disaster Mil Med*. 2015;1(1):14. doi:10.1186/s40696-015-0004-1

60. Essien EJ, Ogungbade GO, Kamiru HN, Ekong E, Ward D, Holmes L Jr. Emerging sociodemographic and lifestyle predictors of intention to use condom in human immunodeficiency virus intervention among uniformed services personnel. *Mil Med*. 2006;171(10):1027-1034. doi:10.7205/MILMED.171.10.1027

61. Faix DJ, Harrison DJ, Riddle MS, et al. Outbreak of Q fever among US military in Western Iraq, June–July 2005. *Clin Infect Dis*. 2008;46(7):e65-e68. doi:10.1086/528866

62. Farrell M, Sebeny P, Klena JD, et al. Influenza risk management: lessons learned from an A(H1N1) pdm09 outbreak investigation in an operational military setting. *PLoS One*. 2013;8(7):e68639. doi:10.1371/journal.pone.0068639

63. Feikin DR, Moroney JF, Talkington DF, et al. An outbreak of acute respiratory disease caused by Mycoplasma pneumoniae and adenovirus at a federal service training academy: new implications from an old scenario. *Clin Infect Dis*. 1999;29(6):1545-1550. doi:10.1086/313500

64. Fernando SD, Booso R, Dharmawardena P, et al. The need for preventive and curative services for malaria when the military is deployed in endemic overseas territories: a case study and lessons learned. *Mil Med Res*. 2017;4(1):19. doi:10.1186/s40779-017-0128-3

65. Frerichs RR, Keim PS, Barrais R, Piarroux R. Nepalese origin of cholera epidemic in Haiti. *Clin Microbiol Infect*. 2012;18(6):E158-E163. doi:10.1111/j.1469-0691.2012.03841.x

66. Fürész J, Lakatos S, Németh K, Fritz P, Simon L, Kacserka K. The prevalence and incidence of Helicobacter pylori infections among young recruits during service in the Hungarian Army. *Helicobacter*. 2004;9(1):77-80. doi:10.1111/j.1083-4389.2004.00200.x

67. Gallimore CI, Pipkin C, Shrimpton H, et al. Detection of multiple enteric virus strains within a foodborne outbreak of gastroenteritis: an indication of the source of contamination. *Epidemiol Infect*. 2005;133(1):41-47. doi:10.1017/S0950268804003218

68. Gambel JM, Drabick JJ, Swalko MA, Henchal EA, Rossi CA, Martinez-Lopez L. Dengue among United Nations mission in Haiti personnel, 1995: implications for preventive medicine. *Mil Med*. 1999;164(4):300-302. doi:10.1093/milmed/164.4.300

69. Gavan DT, Nutt JW. An epidemic of waterborne infectious hepatitis in France. *Arch Environ Health Int J*. 1970;20(4):523-532. doi:10.1080/00039896.1970.10665633

70. Ghose G, Ray KR, Banerjee A. Epidemiological investigation of forest malaria among GREF and army personnel. *Med J Armed Forces India*. 2006;62(1):30-32. doi:10.1016/S0377-1237(06)80149-8

71. Gonzaga VE, Ramos M, Maves RC, Freeman R, Montgomery JM. Concurrent outbreak of norovirus genotype I and enterotoxigenic Escherichia coli on a U.S. Navy ship following a visit to Lima, Peru. *PLoS One*. 2011;6(6):e20822. doi:10.1371/journal.pone.0020822

72. Gray GC, Escamilla J, Hyams KC, Struewing JP, Kaplan EL, Tupponce AK. Hyperendemic Streptococcus pyogenes infection despite prophylaxis with penicillin G benzathine. *N Engl J Med*. 1991;325(2):92-97. doi:10.1056/NEJM199107113250204

73. Gray GC, Goswami PR, Malasig MD, et al. Adult adenovirus infections: loss of orphaned vaccines precipitates military respiratory disease epidemics. *Clin Infect Dis*. 2000;31(3):663-670. doi:10.1086/313999

74. Gremillion DH, Gengler RE, Lathrop GD. Epidemic rubella in military recruits. *South Med J*. 1978;71(8):932-934. doi:10.1097/00007611-197808000-00019

75. Grotto I, Huerta M, Balicer RD, et al. An outbreak of norovirus gastroenteritis on an Israeli military base. *Infection*. 2004;32(6):339-343. doi:10.1007/s15010-004-4002-3

76. Hadad E, Pirogovsky A, Bartal C, et al. An outbreak of leptospirosis among Israeli troops near the Jordan River. *Am J Trop Med Hyg*. 2006;74(1):127-131. doi:10.4269/ajtmh.2006.74.127

77. Halhal B, Glick Y, Galor I, Ran A, Bacon DJ, Glassberg E. Pertussis outbreak among soldiers during basic training: the need for updated protocols. *Mil Med*. 2017;182(S1):355-359. doi:10.7205/MILMED-D-16-00083

78. Hammond-Collins K, Strauss B, Barnes K, et al. Group A streptococcus outbreak in a Canadian Armed Forces training facility. *Mil Med*. 2019;184(3-4):e197-e204. doi:10.1093/milmed/usy198

79. Harbertson J, Grillo M, Zimulinda E, et al. HIV seroprevalence, associated risk behavior, and alcohol use among male Rwanda Defense Forces military personnel. *AIDS Behav*. 2013;17(5):1734-1745. doi:10.1007/s10461-012-0343-6

80. Harbertson J, Scott PT, Moore J, et al. Sexually transmitted infections and sexual behaviour of deploying shipboard US military personnel: a cross-sectional analysis. *Sex Transm Infect*. 2015;91(8):581-588. doi:10.1136/sextrans-2015-052163

81. Harbertson J, Scott PT, Lemus H, Michael NL, Hale BR. Cross-sectional study of sexual behavior, alcohol use, and mental health conditions associated with sexually transmitted infections among deploying shipboard US military personnel. *Mil Med*. 2019;184(11-12):e693-e700. doi:10.1093/milmed/usz070

82. Harris PNA, Oltvolgyi C, Islam A, et al. An outbreak of scrub typhus in military personnel despite protocols for antibiotic prophylaxis: doxycycline resistance excluded by a quantitative PCR-based susceptibility assay. *Microbes Infect*. 2016;18(6):406-411. doi:10.1016/j.micinf.2016.03.006

83. Hart G. Social and psychological aspects of venereal disease in Papua New Guinea. *Br J Vener Dis*. 1974;50(6):453-458. doi:10.1136/sti.50.6.453

84. Hennessy EP. An outbreak of campylobacteriosis amongst directing staff and students at the Infantry Training Centre, Brecon, Wales, March 2004. *J R Army Med Corps*. 2004;150(3):175-178. doi:10.1136/jramc-150-03-03

85. Hernandez RH Jr, Greenberg JH, Olson RE. An outbreak of infectious hepatitis probably due to contamination of food. *Am J Epidemiol*. 1966;84(2):247-252. doi:10.1093/oxfordjournals.aje.a120638

86. Hierholzer JC, Pumarola A, Rodriguez-Torres A, Beltran M. Occurrence of respiratory illness due to an atypical strain of adenovirus type 11 during a large outbreak in Spanish military recruits. *Am J Epidemiol*. 1974;99(6):434-442. doi:10.1093/oxfordjournals.aje.a121632

87. Ho ZJM, Vithia G, Ng CG, et al. Emergence of norovirus GI.2 outbreaks in military camps in Singapore. *Int J Infect Dis*. 2015;31:23-30. doi:10.1016/j.ijid.2014.12.023

88. Hoshino K, Sugiyama M, Date T, et al. Phylogenetic and phylodynamic analyses of hepatitis C virus subtype 1a in Okinawa, Japan. *J Viral Hepat*. 2018;25(8):976-985. doi:10.1111/jvh.12898

89. Huerta M, Grotto I, Gdalevich M, et al. A waterborne outbreak of gastroenteritis in the Golan Heights due to enterotoxigenic Escherichia coli. *Infection*. 2000;28(5):267-271. doi:10.1007/s150100070017

90. Hwang S-M, Park D-E, Yang Y-I, et al. Outbreak of febrile respiratory illness caused by adenovirus at a South Korean military training facility: clinical and radiological characteristics of adenovirus pneumonia. *Jpn J Infect Dis*. 2013;66(5):359-365. doi:10.7883/yoken.66.359

91. Izurieta RO, Macaluso M, Watts DM, et al. Assessing yellow fever risk in the Ecuadorian Amazon. *J Glob Infect Dis*. 2009;1(1):7-13. doi:10.4103/0974-777X.49188

92. Jadhav SL, Sinha AK, Banerjee A, Chawla PS. An outbreak of food poisoning in a military establishment. *Med J Armed Forces India*. 2007;63(2):130-133. doi:10.1016/S0377-1237(07)80055-4

93. Jeger V, Dünki A, Germann M, et al. H1N1 outbreak in a Swiss military boot camp--observations and suggestions. *Swiss Med Wkly*. 2011;141:w13307. doi:10.4414/smw.2011.13307

94. Jelastopulu E, Venieri D, Komninou G, Kolokotronis T, Constantinidis TC, Bantias C. Outbreak of acute gastroenteritis in an air force base in Western Greece. *BMC Public Health*. 2006;6(1):254. doi:10.1186/1471-2458-6-254

95. Jones FR, Ortiz M, Soriano I, et al. Outbreak of gastroenteritis at a Peruvian naval base. *Mil Med*. 2006;171(11):1095-1099. doi:10.7205/MILMED.171.11.1095

96. Kasper MR, Lescano AG, Lucas C, et al. Diarrhea outbreak during U.S. military training in El Salvador. *PLoS One*. 2012;7(7):e40404. doi:10.1371/journal.pone.0040404

97. Kennedy RS. Some notes of an epidemic of dengue form fever amongst Indian troops, Calcutta. *Ind Med Gaz.* 1912;47(11):436-440. https://pubmed.ncbi.nlm.nih.gov/29005395/

98. Khaokham CB, Selent M, Loustalot FV, et al. Seroepidemiologic investigation of an outbreak of pandemic influenza A H1N1 2009 aboard a US Navy vessel-San Diego, 2009. *Influenza Other Respir Viruses*. 2013;7(5):791-798. doi:10.1111/irv.12100

99. Kim K-H, Choi YG, Yoon H-B, et al. Evaluation of the effectiveness of pandemic influenza A(H1N1) 2009 vaccine based on an outbreak investigation during the 2010–2011 season in Korean military camps. *Osong Public Health Res Perspect*. 2013;4(4):209-214. doi:10.1016/j.phrp.2013.07.002

100. Kitsiripornchai S, Markowitz LE, Ungchusak K, et al. Sexual behavior of young men in Thailand: regional differences and evidence of behavior change. *J Acquir Immune Defic Syndr Hum Retrovirol*. 1998;18(3):282-288. doi:10.1097/00042560-199807010-00013

101. Klement E, Uliel L, Engel I, et al. An outbreak of pertussis among young Israeli soldiers. *Epidemiol Infect*. 2003;131(3):1049-1054. doi:10.1017/S0950268803001110

102. Klement E, Talkington DF, Wasserzug O, et al. Identification of risk factors for infection in an outbreak of Mycoplasma pneumoniae respiratory tract disease. *Clin Infect Dis*. 2006;43(10):1239-1245. doi:10.1086/508458

103. Kocik J, Niemcewicz M, Winnicka I, et al. Diversity of influenza-like illness etiology in Polish Armed Forces in influenza epidemic season. *Acta Biochim Pol*. 2014;61(3). doi:10.18388/abp.2014_1869

104. Kolavic‐Gray SA, Binn LN, Sanchez JL, et al. Large epidemic of adenovirus type 4 infection among military trainees: epidemiological, clinical, and laboratory studies. *Clin Infect Dis*. 2002;35(7):808-818. doi:10.1086/342573

105. Konstantinou GN, Papa A, Antoniadis A. Sandfly-fever outbreak in Cyprus: are phleboviruses still a health problem? *Travel Med Infect Dis*. 2007;5(4):239-242. doi:10.1016/j.tmaid.2007.02.002

106. Kotwal RS, Wenzel RB, Sterling RA, et al. An outbreak of malaria in US Army rangers returning from Afghanistan. *JAMA*. 2005;293(2):212-216. doi:10.1001/jama.293.2.212

107. Kuhns DM, Anderson TG. A fly-born bacillary dysentery epidemic in a large military organization. *Am J Public Health Nations Health.* 1944;34(6):750-755. doi:10.2105/ajph.34.7.750

108. Kunwar R, Singh H, Mangla V, Hiremath R. Outbreak investigation: Salmonella food poisoning. *Med J Armed Forces India*. 2013;69(4):388-391. doi:10.1016/j.mjafi.2013.01.005

109. Kunwar R, Sidana N. Mass chemoprophylaxis in control of pneumococcal pneumonia outbreak in a military training centre. *Indian J Public Health*. 2015;59(2):109-114. doi:10.4103/0019-557X.157526

110. Kushwaha AS, Aggarwal SK, Arora MM. Outbreak of meningococcal infection amongst soldiers deployed in operations. *Med J Armed Forces India*. 2010;66(1):4-8. doi:10.1016/S0377-1237(10)80082-6

111. Lagler H, Poeppl W, Winkler H, et al. Hepatitis E virus seroprevalence in Austrian adults: a nationwide cross-sectional study among civilians and military professionals. *PLoS One*. 2014;9(2):e87669. doi:10.1371/journal.pone.0087669

112. Lamar JE 2nd, Malakooti MA. Tuberculosis outbreak investigation of a U.S. Navy amphibious ship crew and the Marine expeditionary unit aboard, 1998. *Mil Med*. 2003;168(7):523-527. https://pubmed.ncbi.nlm.nih.gov/12901459/

113. Laurel VL, De Witt CC, Geddie YA, et al. An outbreak of influenza A caused by imported virus in the United States, July 1999. *Clin Infect Dis*. 2001;32(11):1639-1642. doi:10.1086/320513

114. Lawson R. Observations on the outbreak of yellow fever among the troops at Newcastle, Jamaica, in the latter part of 1856. *Br Foreign Med Chir Rev.* 1859;24(48):445-480. https://www.ncbi.nlm.nih.gov/pmc/articles/PMC5182490/

115. Lee K-J, Kim C-B, Choi B-J, Park K-H, Park J-K. Analysis of vivax malaria cases in Gangwon-do (Province), Korea in the year 2000. *Korean J Parasitol*. 2001;39(4):301-306. doi:10.3347/kjp.2001.39.4.301

116. Lee VJ, Yap J, Tay JK, et al. Seroconversion and asymptomatic infections during oseltamivir prophylaxis against influenza A H1N1 2009. *BMC Infect Dis*. 2010;10(164). doi:10.1186/1471-2334-10-164

117. Leece JD. An outbreak of bacillary dysentery in an isolated army establishment. *BMJ Mil Health.* 1959;105(4):164-167. https://militaryhealth.bmj.com/content/jramc/105/4/164.full.pdf

118. Lesens O, Haus-Cheymol R, Dubrous P, et al. Methicillin-susceptible, doxycycline-resistant Staphylococcus aureus, Côte d’Ivoire. *Emerg Infect Dis*. 2007;13(3):488-490. doi:10.3201/eid1303.060729

119. Lessa FC, Gould PL, Pascoe N, et al. Health care transmission of a newly emergent adenovirus serotype in health care personnel at a military hospital in Texas, 2007. *J Infect Dis*. 2009;200(11):1759-1765. doi:10.1086/647987

120. Letizia AG, Ramos I, Obla A, et al. SARS-CoV-2 transmission among marine recruits during quarantine. *N Eng J Med*. 2020;383(25):2407-2416. doi:10.1056/NEJMoa2029717

121. Levy JW, Bhoomiboonchoo P, Simasathien S, et al. Elevated transmission of upper respiratory illness among new recruits in military barracks in Thailand. *Influenza Other Respir Viruses*. 2015;9(6):308-314. doi:10.1111/irv.12345

122. Lewis T, Loman NJ, Bingle L, et al. High-throughput whole-genome sequencing to dissect the epidemiology of Acinetobacter baumannii isolates from a hospital outbreak. *J Hosp Infect*. 2010;75(1):37-41. doi:10.1016/j.jhin.2010.01.012

123. Li CP. An outbreak of influenza-like disease in the Chinese Army Medical College in 1941. *Am J Public Health Nations Health*. 1958;48(6):760-764. doi:10.2105/AJPH.48.6.760

124. Libório AB, Braz MBM, Seguro AC, et al. Endothelial glycocalyx damage is associated with leptospirosis acute kidney injury. *Am J Trop Med Hyg*. 2015;92(3):611-616. doi:10.4269/ajtmh.14-0232

125. Lichtenstein H. Account of the epidemic dysentery which prevailed among the Dutch troops at the Cape of Good Hope, in 1804 and 1805. *Edinb Med Surg J*. 1810;6(23):296-305. https://www.ncbi.nlm.nih.gov/pmc/articles/PMC5747859/pdf/edinbmedsurgj71835-0036.pdf

126. Lindholm DA, Myers T, Widjaja S, et al. Mosquito exposure and chikungunya and dengue infection among travelers during the chikungunya outbreak in the Americas. *Am J Trop Med Hyg*. 2017;96(4):903-912. doi:10.4269/ajtmh.16-0635

127. Liu W, Kizu JR, Le Grand LR, et al. Localized outbreaks of epidemic polyarthritis among military personnel caused by different sublineages of Ross River virus, Northeastern Australia, 2016–2017. *Emerg Infect Dis.* 2019;25(10):1793-1801. doi:10.3201/eid2510.181610

128. Lopes-João A, Costa I, Mesquita JR, Oleastro M, Penha-Gonçalves C, Nascimento MSJ. Multiple enteropathogenic viruses in a gastroenteritis outbreak in a military exercise of the Portuguese Army. *J Clin Virol*. 2015;68:73-75. doi:10.1016/j.jcv.2015.05.008

129. Lurchachaiwong W, Serichantalergs O, Lertsethtakarn P, et al. Enteric etiological surveillance in acute diarrhea stool of United States military personnel on deployment in Thailand, 2013–2017. *Gut Pathog*. 2020;12(17). doi:10.1186/s13099-020-00356-7

130. MacDonald E, Einöder-Moreno M, Borgen K, et al. National outbreak of Yersinia enterocolitica infections in military and civilian populations associated with consumption of mixed salad, Norway, 2014. *Euro Surveill*. 2016;21(34):30321. doi:10.2807/1560-7917.ES.2016.21.34.30321

131. Mankayi N. Military men and sexual practices: discourses of ‘othering’ in safer sex in the light of HIV/AIDS. *SAHARA J.* 2009;6(1):33-41. doi:10.1080/17290376.2009.9724927

132. Marshall BDL, Prescott MR, Liberzon I, Tamburrino MB, Calabrese JR, Galea S. Posttraumatic stress disorder, depression, and HIV risk behavior among Ohio Army National Guard soldiers: PTSD, depression, and HIV risk in national guard soldiers. *J Trauma Stress*. 2013;26(1):64-70. doi:10.1002/jts.21777

133. McKee KT Jr, Burns WE, Russell LK, et al. Early syphilis in an active duty military population and the surrounding civilian community, 1985–1993. *Mil Med*. 1998;163(6):368-376. doi:10.1093/milmed/163.6.368

134. McNeill KM, Ridgely Benton F, Monteith SC, Tuchscherer MA, Gaydos JC. Epidemic spread of adenovirus type 4-associated acute respiratory disease between U.S. Army installations. *Emerg Infect Dis*. 2000;6(4):415-419. doi:10.3201/eid0604.000419

135. Méndez C, Duque MC, Romero Y, et al. Prevalence of Trypanosoma cruzi infection in active military population of the Colombian National Army gathered in five departments. *PLoS One*. 2019;14(10):e0223611. doi:10.1371/journal.pone.0223611

136. Mgbere O, Monjok E, Abughosh S, Ekong E, Holstad MM, Essien EJ. Modeling covariates of self-perceived and epidemiologic notions of risk for acquiring STIs/HIV among military personnel: a comparative analysis. *AIDS Behav*. 2013;17(3):1159-1175. doi:10.1007/s10461-011-0126-5

137. Michel R, Garnotel E, Spiegel A, Morillon M, Saliou P, Boutin J-P. Outbreak of typhoid fever in vaccinated members of the French Armed Forces in the Ivory Coast. *Eur J Epidemiol*. 2005;20(7):635-642. doi:10.1007/s10654-005-7454-6

138. Mielke JH, Pitkanen KJ. War demography: the impact of the 1808-09 war on the civilian population of Aland, Finland. *Eur J Popul*. 1989;5(4):373-398. doi:10.1007/BF01796793

139. Mitchell RB, Tuttle EE, Dingledine LC, et al. The interpost dissemination of epidemic strains of hemolytic streptococci by troop movements. *J Infect Dis*. 1946;78(2):128-134. doi:10.1093/infdis/78.2.128

140. Mubayi A, Paredes M, Ospina J. A comparative assessment of epidemiologically different cutaneous leishmaniasis outbreaks in Madrid, Spain and Tolima, Colombia: an estimation of the reproduction number via a mathematical model. *Trop Med Infect Dis*. 2018;3(2):43. doi:10.3390/tropicalmed3020043

141. Mukhopadhya J, Kabra SC. Socio behavioural profile of HIV positive DSC personnel. *Med J Armed Forces India*. 2006;62(4):328-331. doi:10.1016/S0377-1237(06)80098-5

142. Mustafa MS, Jain S, Agrawal VK. Food poisoning outbreak in a military establishment. *Med J Armed Forces India*. 2009;65(3):240-243. doi:10.1016/S0377-1237(09)80013-0

143. Myint KSA, Duripunt P, Mammen MP Jr, Sirisopana N, Rodkvamtook W, Gibbons RV. Hepatitis E virus infection in Thai troops deployed with U.N. peacekeeping forces. *Mil Med*. 2007;172(11):1217-1219. doi:10.7205/MILMED.172.11.1217

144. Neela VK, Azhari NN, Joseph N, et al. An outbreak of leptospirosis among reserve military recruits, Hulu Perdik, Malaysia. *Eur J Clin Microbiol Infect Dis*. 2019;38(3):523-528. doi:10.1007/s10096-018-03450-6

145. Neo FJX, Loh JJP, Ting P, et al. Outbreak of caliciviruses in the Singapore military, 2015. *BMC Infect Dis*. 2017;17(1):719. doi:10.1186/s12879-017-2821-y

146. Nivesvivat T, Janthayanont D, Mungthin M, et al. Methicillin-susceptible Staphylococcus aureus skin infections among military conscripts undergoing basic training in Bangkok, Thailand, in 2014. *BMC Res Notes*. 2016;9(179). doi:10.1186/s13104-016-1989-3

147. Nowosiwsky T. The epidemic curve of Plasmodium falciparum malaria in a nonimmune population. American troops in Vietnam, 1965 and 1966. *Am J Epidemiol*. 1967;86(2):461-467. doi:10.1093/oxfordjournals.aje.a120756

148. Nwokoji UA, Ajuwon AJ. Knowledge of AIDS and HIV risk-related sexual behavior among Nigerian naval personnel. *BMC Public Health*. 2004;4(24). doi:10.1186/1471-2458-4-24

149. Ochiai H, Ohtsu T, Tsuda T, et al. Clostridium perfringens foodborne outbreak due to braised chop suey supplied by chafing dish. *Acta Med Okayama*. 2005;59(1):27-32. doi:10.18926/AMO/31986

150. Okeke CE, Onwasigwe CN, Ibegbu MD. The effect of age on knowledge of HIV/AIDS and risk related behaviours among army personnel. *Afr Health Sci*. 2012;12(3):291-296. doi:10.4314/ahs.v12i3.7

151. Oré M, Sáenz E, Cabrera R, et al. Outbreak of cutaneous leishmaniasis in Peruvian military personnel undertaking training activities in the Amazon basin, 2010. *Am J Trop Med Hyg*. 2015;93(2):340-346. doi:10.4269/ajtmh.15-0107

152. Oristo S, Rönnqvist M, Aho M, et al. Contamination by norovirus and adenovirus on environmental surfaces and in hands of conscripts in two Finnish garrisons. *Food Environ Virol*. 2017;9(1):62-71. doi:10.1007/s12560-016-9262-4

153. Pang J, Jin J, Loh JP, et al. Risk factors for febrile respiratory illness and mono-viral infections in a semi-closed military environment: a case-control study. *BMC Infect Dis*. 2015;15(288). doi:10.1186/s12879-015-1024-7

154. Papaevangelou G, Mendris J, Kyriakidou A. Rubella epidemic in a naval training center. *Am J Epidemiol*. 1969;89(6):665-668. doi:10.1093/oxfordjournals.aje.a120980

155. Paul JR, McClure WW. Epidemic hemorrhagic fever attack rates among United Nations troops during the Korean war. *Am J Hyg*. 1958;68(2):126-139. doi:10.1093/oxfordjournals.aje.a119957

156. Peczenik A, Duttweiler DW, Moser RH. An apparently water-borne outbreak of infectious hepatitis. *Am J Public Health Nations Health*. 1956;46(8):1008-1017. doi:10.2105/AJPH.46.8.1008

157. Petersen LR, Sawyer LA, Fishbein DB, et al. An outbreak of ehrlichiosis in members of an army reserve unit exposed to ticks. *J Infect Dis*. 1989;159(3):562-568. doi:10.1093/infdis/159.3.562

158. Pirnay J-P, Selhorst P, Cochez C, et al. Study of a SARS-CoV-2 outbreak in a Belgian military education and training center in Maradi, Niger. *Viruses*. 2020;12(9):949. doi:10.3390/v12090949

159. Pon E, McKee KT Jr, Diniega BM, Merrell B, Corwin A, Ksiazek TG. Outbreak of hemorrhagic fever with renal syndrome among U.S. Marines in Korea. *Am J Trop Med Hyg*. 1990;42(6):612-619. doi:10.4269/ajtmh.1990.42.612

160. Prier RE, Cowan DN. Risk factors for hepatitis B virus infection in US Army soldiers in Europe. *J Epidemiol Community Health*. 1987;41(3):229-232. doi:10.1136/jech.41.3.229

161. Qu F, Cui E, Guo T, et al. Nasal colonization of and clonal transmission of methicillin-susceptible Staphylococcus aureus among Chinese military volunteers. *J Clin Microbiol*. 2010;48(1):64-69. doi:10.1128/JCM.01572-09

162. Riddle MS, Smoak BL, Thornton SA, Bresee JS, Faix DJ, Putnam SD. Epidemic infectious gastrointestinal illness aboard U.S. Navy ships deployed to the Middle East during peacetime operations – 2000–2001. *BMC Gastroenterol*. 2006;6(9). doi:10.1186/1471-230X-6-9

163. Royal J, Riddle MS, Mohareb E, Monteville MR, Porter CK, Faix DJ,. Seroepidemiologic survey for coxiella burnetii among US military personnel deployed to Southwest and Central Asia in 2005. *Am J Trop Med Hyg*. 2013;89(5):991-995. doi:10.4269/ajtmh.12-0174

164. Russell KL, Gonzalez MAM, Watts DM, et al. An outbreak of leptospirosis among Peruvian military recruits. *Am J Trop Med Hyg*. 2003;69(1):53-57. doi:10.4269/ajtmh.2003.69.53

165. Ryan MAK, Gray GC, Smith B, McKeehan JA, Hawksworth AW, Malasig MD. Large epidemic of respiratory illness due to adenovirus types 7 and 3 in healthy young adults. *Clin Infect Dis.* 2002;34(5):577-582. doi:10.1086/338471

166. Sanchez JL, Bendet I, Max Grogl L, et al. Malaria in Brazilian military personnel deployed to Angola. *J Travel Med*. 2000;7(5):275-282. doi:10.2310/7060.2000.00077

167. Sanchez JL, Binn LN, Innis BL, et al. Epidemic of adenovirus-induced respiratory illness among US military recruits: epidemiologic and immunologic risk factors in healthy, young adults. *J Med Virol*. 2001;65(4):710-718. doi:10.1002/jmv.2095

168. Schmid D, Kasper S, Kuo HW, et al. Ongoing rubella outbreak in Austria, 2008-2009. *Euro Surveill*. 2009;14(16):19184. doi:10.2807/ese.14.16.19184-en

169. Seah SG-K, Lim EA-S, Kok-Yong S, et al. Viral agents responsible for febrile respiratory illnesses among military recruits training in tropical Singapore. *J Clin Virol*. 2010;47(3):289-292. doi:10.1016/j.jcv.2009.12.011

170. Sebeny PJ, Pimentel G, Cline J, et al. Hotel clinic-based diarrheal and respiratory disease surveillance in U.S. service members participating in operation bright star in Egypt, 2009. *Am J Trop Med Hyg*. 2012;87(2):312-318. doi:10.4269/ajtmh.2012.11-0318

171. Seña AC, Miller WC, Hoffman IF, et al. Trends of gonorrhea and chlamydial infection during 1985–1996 among active-duty soldiers at a United States Army installation. *Clin Infect Dis*. 2000;30(4):742-748. doi:10.1086/313742

172. Sharma S, Sahu R, Taneja NS, Hazra A. Epidemiological investigation of viral hepatitis E outbreak in two colocated military training centers, North India, April–June 2016. *Med J Armed Forces India*. 2019. doi:10.1016/j.mjafi.2018.09.001

173. Sharp TW, Hyams KC, Watts D, et al. Epidemiology of Norwalk virus during an outbreak of acute gastroenteritis aboard a US aircraft carrier. *J Med Virol*. 1995;45(1):61-67. doi:10.1002/jmv.1890450112

174. Singh M, Kalghatgi A, Narayanan K, Rao K, Nagendra A. Outbreak of salmonella food poisoning at high altitude. *Med J Armed Forces India*. 1998;54(2):96-98. doi:10.1016/S0377-1237(17)30490-2

175. Singh P, Handa SK, Banerjee A RETD. Epidemiological investigation of an outbreak of viral hepatitis. *Med J Armed Forces India*. 2006;62(4):332-334. doi:10.1016/S0377-1237(06)80100-0

176. Steinberg P, White RJ, Fuld SL, Gutekunst RR, Chanock RM, Sentherfit LB. Ecology of Mycoplasma pneumoniae infections in marine recruits at Parris Island, South Carolina. *Am J Epidemiol*. 1969;89(1):62-73. doi:10.1093/oxfordjournals.aje.a120916

177. Struewing JP, Gray GC. An epidemic of respiratory complaints exacerbated by mass psychogenic illness in a military recruit population. *Am J Epidemiol*. 1990;132(6):1120-1129. doi:10.1093/oxfordjournals.aje.a115755

178. Sundnes KO, Haimanot, AT. Epidemic of louse-borne relapsing fever in Ethiopia. *The Lancet*. 1993;342(8881):1213-1215. doi:10.1016/0140-6736(93)92190-5

179. Supramaniam V, Tan DS. An outbreak of rubella among Malaysian Air Force recruits, 1979. *Med J Malaysia.* 1980;34(4):415-417. https://pubmed.ncbi.nlm.nih.gov/7219274/

180. Suryam V, Bhatti VK, Kulkarni A, Mahen A, Nair V. Outbreak control of community acquired pneumonia in a large military training institution. *Med J Armed Forces India*. 2015;71(1):33-37. doi:10.1016/j.mjafi.2014.09.015

181. Suwannahitatorn P, Jatapai A, Rangsin R. An outbreak of Paederus dermatitis in Thai military personnel. *J Med Assoc Thai.* 2014;97 Suppl 2:S96-S100. https://pubmed.ncbi.nlm.nih.gov/25518181/

182. Szwarcwald CL, de Carvalho MF, Barbosa Júnior A, Barreira D, Speranza FAB, de Castilho EA. Temporal trends of HIV-related risk behavior among Brazilian military conscripts, 1997-2002. *Clinics*. 2005;60(5):367-374. doi:10.1590/S1807-59322005000500004

183. Taamasri P, Leelayoova S, Rangsin R, Naaglor T, Ketupanya A, Mungthin M. Prevalence of Blastocystis hominis carriage in Thai Army personnel based in Chonburi, Thailand. *Mil Med*. 2002;167(8):643-646. doi:10.1093/milmed/167.8.643

184. Tam CC, Anderson KB, Offeddu V, et al. Epidemiology and transmission of respiratory infections in Thai army recruits: a prospective cohort study. *Am J Trop Med Hyg*. 2018;99(4):1089-1095. doi:10.4269/ajtmh.18-0219

185. Tansel O, Ekuklu G, Otkun M, Otkun MT, Akata F, Tuğrul M. A food-borne outbreak caused by Salmonella enteritidis. *Yonsei Med J*. 2003;44(2):198-202. doi:10.3349/ymj.2003.44.2.198

186. Tarabbo M, Lapa D, Castilletti C, et al. Retrospective investigation of an influenza A/H1N1pdm outbreak in an Italian military ship cruising in the Mediterranean Sea, May-September 2009. *PLoS One*. 2011;6(1):e15933. doi:10.1371/journal.pone.0015933

187. Tate JE, Bunning ML, Lott L, et al. Outbreak of severe respiratory disease associated with emergent human adenovirus serotype 14 at a US air force training facility in 2007. *J Infect Dis*. 2009;199(10):1419-1426. doi:10.1086/598520

188. Theamboonlers A, Rianthavorn P, Jiamsiri S, et al. Molecular characterization of hepatitis A virus causing an outbreak among Thai Navy recruits. *Trop Biomed*. 2009;26(3):352-359.

189. Thornton S, Davies D, Chapman F, et al. Detection of Norwalk-like virus infection aboard two U.S. Navy ships. *Mil Med*. 2002;167(10):826-830. https://pubmed.ncbi.nlm.nih.gov/12392249/

190. Thornton SA, Sherman SS, Farkas T, Zhong W, Torres P, Jiang X. Gastroenteritis in US Marines during Operation Iraqi Freedom. *Clin Infect Dis*. 2005;40(4):519-525. doi:10.1086/427501

191. Todkill D, Pudney R, Terrell A, et al. An outbreak of Shigella boydii serotype 20 in January 2015 amongst United Kingdom healthcare workers involved in the Ebola response in Sierra Leone. *J Med Microbiol*. 2018;67(11):1596-1600. doi:10.1099/jmm.0.000832

192. Tong C, Javelle E, Grard G, et al. Tracking rift valley fever: from Mali to Europe and other countries, 2016. *Euro Surveill*. 2019;24(8):1800213. doi:10.2807/1560-7917.ES.2019.24.8.1800213

193. Torres-Slimming PA, Mundaca, CC, Moran M, et al. Outbreak of cyclosporiasis at a naval base in Lima, Peru. *Am J Trop Med Hyg*. 2006;75(3):546-548. doi:10.4269/ajtmh.2006.75.546

194. Tran BR, Davis A, Ito SI, et al. Alcohol and cannabis use and sexual risk behaviors in the Malawi Defence Force. *AIDS Behav*. 2018;22(9):2851-2860. doi:10.1007/s10461-018-2167-5

195. Tuck JJH, Williams JR, Doyle AL. Gastro enteritis in a military population deployed in West Africa in the UK Ebola response; was the observed lower disease burden due to handwashing? *Travel Med Infect Dis*. 2016;14(2):131-136. doi:10.1016/j.tmaid.2015.12.009

196. Vainio A, Lyytikäinen O, Sihvonen R, et al. An outbreak of pneumonia associated with S. pneumoniae at a military training facility in Finland in 2006. *APMIS*. 2009;117(7):488-491. doi:10.1111/j.1600-0463.2009.02463.x

197. Vickery JP, Tribble DR, Putnam SD, et al. Factors associated with the use of protective measures against vector-borne diseases among troops deployed to Iraq and Afghanistan. *Mil Med*. 2008;173(11):1060-1067. doi:10.7205/MILMED.173.11.1060

198. Wadl M, Scherer K, Nielsen S, et al. Food-borne norovirus-outbreak at a military base, Germany, 2009. *BMC Infect Dis*. 2009;10(30). doi:10.1186/1471-2334-10-30

199. Wallace MR, Sharp TW, Smoak B, et al. Malaria among United States troops in Somalia. *Am J Med*. 1996;100(1):49-55. doi:10.1016/S0002-9343(96)90011-X

200. Walton BC, Person DA, Bernstein R. Leishmaniasis in the U.S. military in the Canal Zone. *Am J Trop Med Hyg*. 1968;17(1):19-24. doi:10.4269/ajtmh.1968.17.19

201. Warner RD, Carr RW, McCleskey FK, Johnson PC, Elmer LM, Davison VE. A large nontypical outbreak of Norwalk virus. Gastroenteritis associated with exposing celery to nonpotable water and with Citrobacter freundii. *Arch Intern Med*. 1991;151(12):2419-2424. doi:10.1001/archinte.151.12.2419

202. Watier-Grillot S, Boni M, Tong C, et al. Challenging investigation of a norovirus foodborne disease outbreak during a military deployment in Central African Republic. *Food Environ Virol*. 2017;9(4):498-501. doi:10.1007/s12560-017-9312-6

203. Wentz LM, Ward MD, Potter C, et al. Increased risk of upper respiratory infection in military recruits who report sleeping less than 6 h per night. *Mil Med*. 2018;183(11-12):e699-e704. doi:10.1093/milmed/usy090

204. Witkop CT, Duffy MR, Macias EA, et al. Novel influenza A (H1N1) outbreak at the U.S. Air Force Academy: epidemiology and viral shedding duration. *Am J Prev Med*. 2010;38(2):121-126. doi:10.1016/j.amepre.2009.10.005

205. Yagupsky P, Ben-Ami Y, Trefler R, Porat N. Outbreaks of invasive Kingella kingae infections in closed communities. *J Pediatr*. 2016;169:135-139.e1. doi:10.1016/j.jpeds.2015.10.025

206. Yap J, Qadir A, Liu I, Loh J, Tang BH, Lee VJ. Outbreak of acute norovirus gastroenteritis in a military facility in Singapore: a public health perspective. *Singapore Med J.* 2012;53(4):249-254. https://pubmed.ncbi.nlm.nih.gov/22511047/

207. Yoon CG, Kang DY, Jung J, et al. The infectivity of pulmonary tuberculosis in Korean army units: evidence from outbreak investigations. *Tuberc Respir Dis (Seoul)*. 2019;82(4):298-305. doi:10.4046/trd.2018.0077

208. Yu P, Ma C, Nawaz M, et al. Outbreak of acute respiratory disease caused by human adenovirus type 7 in a military training camp in Shaanxi, China. *Microbiol Immunol*. 2013;57(8):553-560. doi:10.1111/1348-0421.12074

209. Ziebold C, Hassenpflug B, Wegner-Bröse H, Wegner K, Schmitt HJ. An outbreak of rubella aboard a ship of the German Navy. *Infection*. 2003;31(3):136-142. doi:10.1007/s15010-003-3048-y

210. Zinderman CE, Conner B, Malakooti MA, LaMar JE, Armstrong A, Bohnker BK. Community-acquired Methicillin-resistant Staphylococcus aureus among military recruits. *Emerg Infect Dis*. 2004;10(5):941-944. doi: 10.3201/eid1005.030604
